# A foundation model for generalized brain MRI analysis

**DOI:** 10.1101/2024.12.02.24317992

**Authors:** Divyanshu Tak, Biniam A. Garomsa, Tafadzwa L. Chaunzwa, Anna Zapaishchykova, Juan Carlos Climent Pardo, Zezhong Ye, John Zielke, Yashwanth Ravipati, Sri Vajapeyam, Maryam Mahootiha, Ceilidh Smith, Ariana M. Familiar, Kevin X. Liu, Sanjay Prabhu, Pratiti Bandopadhayay, Ali Nabavizadeh, Sabine Mueller, Hugo JWL Aerts, Raymond Y. Huang, Tina Y. Poussaint, Benjamin H. Kann

**Affiliations:** Artificial Intelligence in Medicine (AIM) Program, Mass General Brigham, Harvard Medical School, Boston, MA, United States; Department of Radiation Oncology, Dana-Farber Cancer Institute and Brigham and Women’s Hospital, Harvard Medical School, Boston, MA, United States; Radiology and Nuclear Medicine, CARIM & GROW, Maastricht University, Maastricht, the Netherlands; Boston Children’s Hospital, Boston, MA, United States; Dana-Farber Cancer Institute, Boston, MA, United States; Michigan State University, East Lansing, MI, United States; Children’s Hospital of Philadelphia, Philadelphia, United States; University of Pennsylvania, Pennsylvania, United States; Department of Neurology, Neurosurgery and Pediatrics, University of California, San Francisco, United States; Memorial Sloan Kettering Cancer Center, New York, United States; Department of Radiology, Brigham and Women’s Hospital, Harvard Medical School, Boston MA

**Keywords:** Self-supervised learning, Deep-Learning, Foundation Model, Brain MRI, Artificial Intelligence

## Abstract

Artificial intelligence (AI) applied to brain magnetic resonance imaging (MRI) has the potential to improve disease diagnosis and management but requires algorithms with generalizable knowledge that can perform well in a variety of clinical scenarios. The field has been constrained, thus far, by limited training data and task-specific models that do not generalize well across patient populations and medical tasks. Foundation models, by leveraging self-supervised learning, pretraining, and targeted adaptation, present a promising paradigm to overcome these limitations. Here, we present Brain Imaging Adaptive Core (BrainIAC), a novel foundation model designed to learn generalized representations from unlabeled brain MRI data and serve as a core basis for diverse downstream application adaptation. Trained and validated on 48,519 brain MRIs across a broad spectrum of tasks, we demonstrate that BrainIAC outperforms localized supervised training and other pretrained models, particularly in low-data settings and high-difficulty tasks, allowing for application in scenarios otherwise infeasible. BrainIAC can be integrated into imaging pipelines and multimodal frameworks and may lead to improved biomarker discovery and AI clinical translation.

## INTRODUCTION

Recent advances in deep learning have transformed medical artificial intelligence (AI), enabling the development of clinically translatable tools that, in some clinical tasks, match or even surpass expert performance ^1,2^. Medical practice, with its vast and diverse data sources across different modalities—clinical notes, histopathology images, radiographic images, and genomics—presents a compelling landscape for applied AI to synthesize data, learn patterns, and make predictions. However, the scarcity and heterogeneity of labeled data, particularly for rare diseases and cases involving expensive data acquisition procedures, such as brain magnetic resonance imaging (MRI), remains a significant barrier to the development of clinically useful AI imaging tools.

Self-supervised learning (SSL) has emerged as a promising advance to traditional supervised learning methods, with its ability to learn inherent, generalizable information from large unlabeled data that are much more available than annotated, task-specific dataset. This approach allows for the extraction of meaningful representations from unlabeled data, that can be easily transferred to different applications. SSL methods have demonstrated remarkable success in computer vision ^3,4,5,6^ and natural language processing ^7,8^, with recent translations into medicine ^9–13^. This shift has facilitated a transition from narrow-task learning of medical AI models to a more generalized task agnostic learning coupled with localized fine-tuning. The resulting algorithms, often referred to as foundation models, have shown substantial potential in developing clinically employable solutions across various medical domains ^14–16^.

Despite these advancements, however, the application of SSL to 3D brain MRI has been limited. The high-dimensional, heterogeneous nature of brain MRI data presents unique challenges to the development of performant models. Unlike other imaging modalities such as computed tomography (CT), brain MRI has variety of acquisition sequences from a single scan that vary by institution and scanner, with classic examples including T1-weighted, T2-weighted, and T1-weighted with gadolinium contrast enhancement (T1CE) —each providing distinct sets of information, with selection of sequences for analysis depending upon the clinical use cases. For example, T1-weighted sequences are commonplace for neurocognitive analysis of pediatric and adolescents as well as later neurofunctional diseases (e.g. Alzheimer’s dementia)^17,18^.

Whereas T2-weighted sequences are preferred for lesion segmentation^1,19^, compared to T2-fluid attenuation inversion recovery (FLAIR) and T1CE, which are commonly used for response assessment for brain tumors^20,21^. Variability in MRI scanner, acquisition protocol, and patient setup also introduce biases in voxel intensities, which are problematic for radiomic analyses^22,23^. Furthermore, MR acquisition itself is subject to variability and noise, with a range of scanners (from 1.5T to 7T) and differing acquisition parameters (echo time, relaxation time)^24^. Foundation models for MRI must overcome substantial heterogeneity in brain structural features across different age groups and (sometimes rare) disease pathologies, which may constrain generalizable feature extraction.

Prior investigations have proposed foundation model frameworks for brain lesion segmentation^25^ and aging-related tasks ^26^, but there remains a need for a broadly generalizable model for both healthy and abnormal brain images. In this study, we introduce a brain imaging adaptive core (BrainIAC), a general, multiparametric brain MRI foundation model based on SSL principles. Developed and validated in 48,519 brain MRIs with a wide spectrum of scanner parameters, demographics, and medical settings, we show that BrainIAC learns robust and adaptable representations. We evaluate BrainIAC performance on multiple downstream applications across a range of clinical settings with varying task complexity. We compare BrainIAC to traditional supervised learning approaches and transfer learning from pretrained medical imaging networks^27^. We perform stability analysis to simulate real world acquisition and demographic variance and compare the robustness and generalizability of the BrainIAC learned features to other approaches. Our findings demonstrate BrainIAC’s versatility and ability to adapt to multiple clinical settings, providing a usable foundation to accelerate computational brain imaging analysis research.

## RESULTS

We pretrained BrainIAC using self-supervised contrastive learning on 32,000 multiparametric MRIs curated from 35 datasets across ten medical conditions (Figure 1; Supplementary Data Table 1-4). We show that BrainIAC is adaptable to four distinct, clinically meaningful, downstream prediction tasks, outperforming current supervised and transfer learning approaches, and that successful adaptation requires only limited data for fine-tuning. The four tasks are MRI sequence classification, brain age prediction, isocitrate dehydrogenase (IDH) mutation detection, and survival prediction for brain tumors. These tasks were chosen as they represented a wide range of difficulty (e.g. MRI sequence classification is straightforward for a human clinician, while mutational status prediction is extremely challenging) and clinical contexts. For each downstream application we compared BrainIAC to localized supervised training (Scratch), and a medical-imaging specific model (MedicalNet^27^). We compared the performance across limited data scenarios, increasing fraction of finetuning data available from 10% to 100% with independent test sets for performance metrics (Extended Data Fig.1). Finally, we analyzed resiliency of BrainIAC and benchmark models to image-related artifacts.

**Figure 1.**
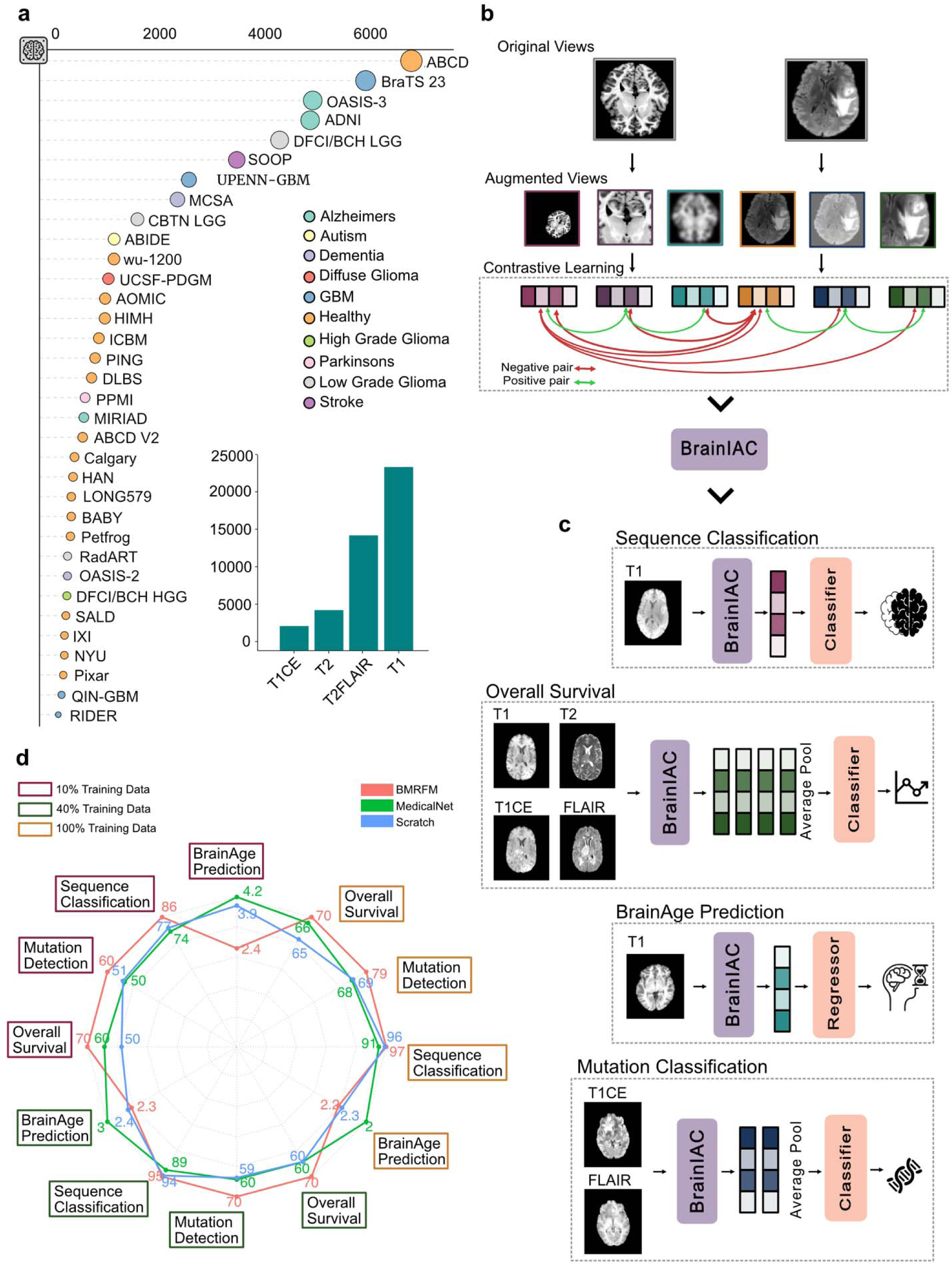
Overview of the study. BrainIAC is a general-purpose foundation model for Brain MR imaging analysis, trained using self-supervised contrastive learning approach and validated on 4 diverse downstream applications, BrainIAC outuperforms supervised training (Scratch), and finetuning from available pretrained model (MedicalNet). BrainIAC serves as a vision encoder for 3D Brain MRs generating robust latent feature representation which can be easily adapted to downstream applications. **A** Datasets used in the study, a pool of 35 datasets ranging across 10 neurological conditions and 4 sequences totalling upto 48,519 brain MRI images was curated and preprocessed. **B** BrainIAC was trained using contrastive learning based self-supervised learning approach SimCLR on 32,000 Brain MR images, which includes loss minimization by positive pair attraction and negative pair repulsion. **C** BrainIAC was evaluated in 4 downstream settings : Sequence classification, BrainAge Prediction, Mutation Classification, and Overall Survival. Each application leveraged BrainIAC as the vision encoder with task-specific architectures. **D** BrainIAC outperforms other approaches (Scratch, MedicalNet) for downstream application at varying data availability settings (10%, 40%, 100%).

### MRI sequence Classification

Sequence classification is a critical, upstream step in MRI curation and processing that remains difficult given heterogeneity in scanner protocols and inconsistent documentation of sequence details at acquisition. While deep learning has shown promise in automated classification, there remains room for improvement, particularly in the classification of contrast enhancement in T1-weight scans^28^. We utilized 5,005 scans for fine-tuning (or in the case of the Scratch model, training) and validation of all three approaches, with a reserved holdout set of 876 scans encompassing the four primary sequences used in brain tumor assessment (T1, T2, T1CE, FLAIR) from the BraTS 2023 dataset^29^ (Supplementary Data Table 5). We found that performance increased incrementally with fine-tuning data availability. At lower data availability, BrainIAC outperformed MedicalNet and Scratch models (Figure 2a). For example, at 10% availability (n=500 scans), BrainIAC balanced accuracy (BA) was 86.4%, MedicalNet was 74.2%, and Scratch was 79.0%. BrainIAC continued to outperform other models until 60% (n=3000) of scans were available for training, at which point performance plateaued for all models (BrainIAC BA: 97.1%, MedicalNet BA: 93.4%, Scratch BA: 96.3%). We performed KNN clustering (K=4, representing each sequence) on the features and calculated the Davies-Bouldin Index scores. For 100% data BrainIAC demonstrated better clustering performance with Davies-Bouldin Index of 0.68 (Figure 2b) compared to 0.72 of Scratch and 0.82 of MedicalNet (Supplementary Data Table 6).

**Figure 2.**
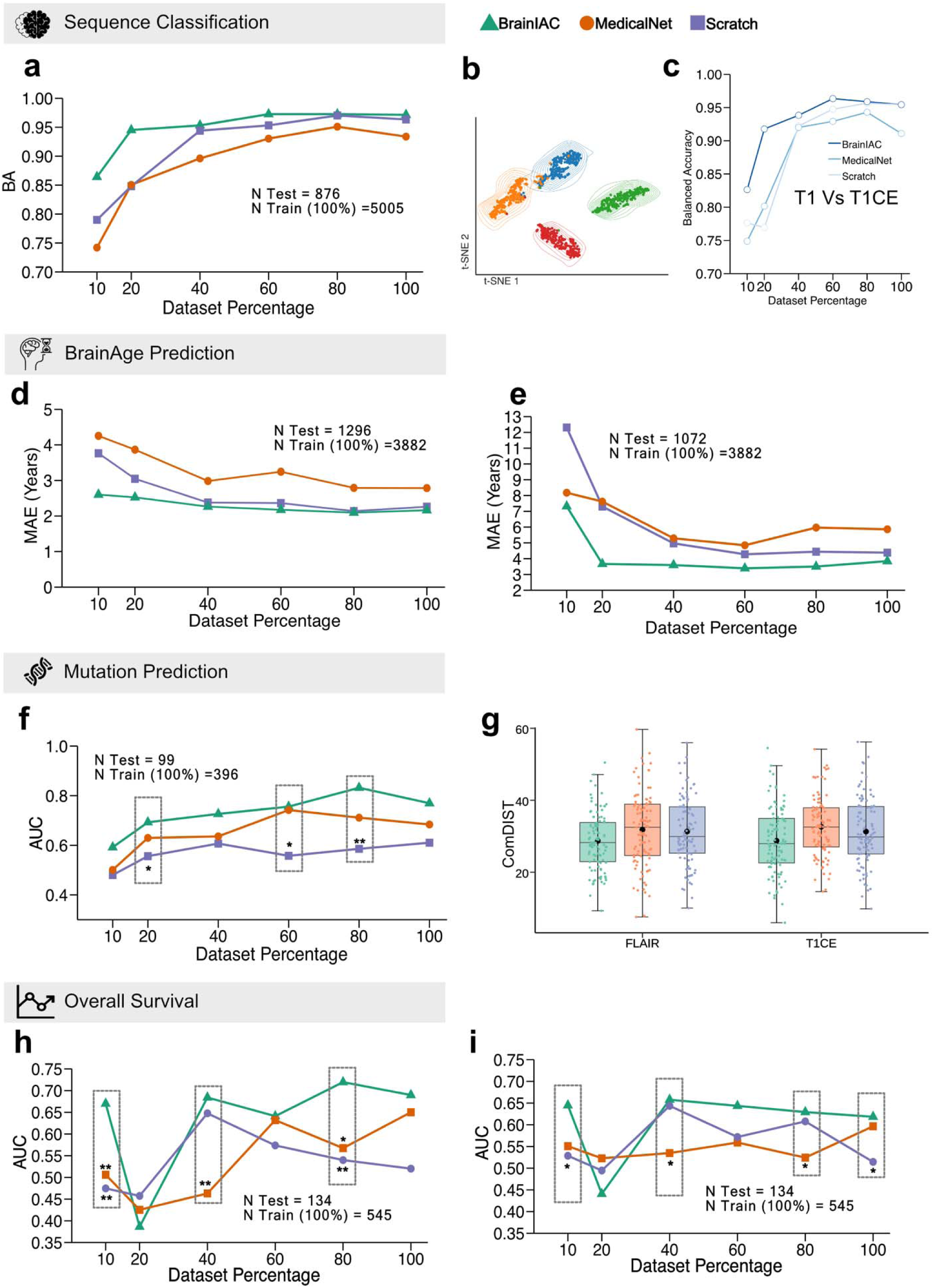
Downstream application performance comparison. **A** Sequence Classification. Line plot depicts the performance (balanced accuracy) comparison of BrainIAC, Scratch, MedicalNet for to perform 4-way classification between T1, T2, T1CE, and T2FLAIR input sequences. The total images used for finetuning with 100% data is 5005 with n=4000 (80%), n=3000 (60%), n=2000 (40), n=1000 (20%), n=500(10%), with 876 images reserved as holdout set for testing. ***B*** depicts the latent feature representation of the BrainIAC finetund model on the test set with contour overlay. KNN clustering was performed on the latent features for of the BrainIAC finetuned model with Davies-Bouldin score of 0.68**. C** displays the subgroup analysis on T1 Vs T1CE images for all 3 approaches across all data fractions. **D** BrainAge prediction. The line plot depicts the performance(MAE) comparison of BrainIAC with Scratch and MedicalNet on the In-distribution test set (n=1296). ***D*** Line plot shows the performance comparison of three approaches on out-of-distribution test set (n=1072). Training for all three aproaches was done on total 3882 images (100%) with n=3100 (80%), n=2325 (60%), n=1550 (40), n=777 (20%), n=388(10%). **F.** Mutation prediction. The line plot left depicts the performance comparison of the three approaches for IDH Mutation classification (IDH mutated Vs Wildtype) on a holdout set of 99 scans. The training for all 3 approaches was performed on 396 scans(100%), n=316 (80%), n=237 (60%), n=158 (40), n=80 (20%), n=40(10%). ***G***. The boxplot depicts the ComDIST scores of the the BrainIAC, Scratch and MedicalNet trained on 100% data (n=396). **H** Overall survival. The left line plots depicts the AUC comparison of 1year overall survival of GBM subjects for the three approaches on a holdout set of 134 subjects. The trainings for all approaches were performed with n=534 (100%), n=427 (80%), n=320 (60%), n=214 (40%), n=107 (20%), n=53(10%). ***I***. The line plot depicts the AUC comparison of 1year overall survival of GBM subjects for the three approaches on an external test set of 134 subjects.

Subgroup analysis of T1CE versus T1 classification, which is seen as the most challenging identification task, demonstrated a similar trend, with BrainIAC outperforming the Scratch and MedicalNet at training data availability below 80% (Figure 2c). Consistent findings were observed in additional subgroup analyses, including T2 vs. FLAIR, T1 vs. T2, and FLAIR vs. T1CE (Extended Data Fig.2).

### Brain age Prediction

MRI-based brain age prediction is associated with neurocognitive function in aging adults and may have utility as an early biomarker for Alzheimer’s disease^30,31^. To evaluate BrainIAC as a foundation for improved brain age prediction, we aggregated a dataset of 6,250 T1-weighted scans, allocating 3,882 for training/validation and 1,296 as an in-distribution, internal test set. and an out-of-distribution test set of 1,072 scans from ABCD V2, IXI, Long579, and Pixar datasets (Supplementary Data Table 7,8). Performance was assessed using Mean Absolute Error (MAE) for predicted age versus chronological age.

In both internal and external test sets, performance improved with training data availability, and BrainIAC outperformed other models at lower data availability (Figure 2d,2e). In the external test set, at 20% training data availability (n=775 scans), BrainIAC achieved MAE of 3.95 (95% CI: 3.78 -4.13), compared to MedicalNet MAE of 7.61 years (95% CI: 7.35-7.87), and Scratch MAE of 6.89 years (95% CI: 6.63-7.16) (p<0.0001). BrainIAC continued to outperform other models until 100% of training data was available. Findings were consistent in subgroup analyses on the external test sets by individual data source (Figure 5e,5f).

BrainIAC demonstrated increased accuracy at all age predictions compared with MedicalNet and Scratch models with a reduced delta between the predicted age and chronological age (Figure 5a, 5b). The t-SNE representations of BrainIAC demonstrate clear clustering of the latent features based on the age groups: 0-10 years, 10-20 years, 20-30 years, 30-40 years (Figure 5c, 5d). With a Davies-Boulding index score of 0.575 (External) and 0.475 (Internal) BrainIAC outperforms MedicalNet 0.633 (External) 0.538 (Interna) and Scratch 0.612 (External) and 0.485 (Internal) on clustering of age binned latent features.

### Cancer mutational subtype prediction

Non-invasive, imaging-based prediction of brain tumor mutational subtypes could provide actionable information to dictate clinical management when tissue biopsy is deemed infeasible^32–34^. We evaluated BrainIAC as a foundation for tumor mutational subtyping IDH mutation prediction in low-grade glioma setting. Performance was evaluated using the area under the receiver operating characteristic curve (AUC) for discriminating mutation versus wildtype tumor (Figure 2f). The DeLong^35^ test was used for checking statistical significance and calculating P-Values.

For the IDH classification, we utilized 396 scans for training/validation and 99 reserved as a test set from the UCSF-PDGM^36^ dataset (Supplementary Data Table 9). Performance increased incrementally with training data availability, but, in this case of very limited data, BrainIAC consistently outperformed other models at all levels of data availability. At 10% data availability (n=50), BrainIAC yielded AUC 0.60 (95% CI: 0.45-0.72), compared to 0.50 (95% CI: 0.36-0.63) for MedicalNet and 0.49 (95% CI: 0.34-0.62) for Scratch. Increasing training data availability to 100% (n=396), BrainIAC yielded AUC 0.76 (95% CI: 0.61-0.90), compared to 0.68 (95% CI: 0.54-0.81) for MedicalNet, and 0.61 (95% CI: 0.48-0.73) for Scratch (p=0.014) (Supplementary Data Table 10).

We generated saliency maps for BrainIAC to visualize the model’s internal weight activation and attention (Figure 6c). We quantified the model attention with respect to tumor region using the ComDIST^32^ analysis. The ComDIST scores revealed a closer attention of BrainIAC to tumor region of interest with score of 28.70 (T1CE) and 28.73(FLAIR) compared to MedicalNet 31.82 (T1CE) and 32.60(FLAIR) and Scratch 31.29 (T1CE) and 31.32(FLAIR) (Figure 2g) (Supplementary Data Table 11).

### Overall Survival prediction

Computational analysis of cancer imaging data can potentially improve prognostication and risk-stratification beyond traditional staging^37^. We evaluated BrainIAC as a foundation for survival prediction for glioblastoma multiforme (GBM) using the UPENN-GBM^38^ dataset. Of 671 patients total, 668 patients with complete survival information were included (Supplementary Data Table 12). We randomly split the dataset with 534 (80%) patients for fine-tuning and 134 (20%) reserved as a test set. We further performed external testing on 134 patients from TCGA-GBM/Brats23 dataset with complete survival information. Model performance was assessed using AUC for predicting survival at 1year post-treatment with complete data up through this timepoint. Median model risk score output was used to stratify patients into low-and high-risk groups with Kaplan-Meier survival curves calculated and log-rank tests to compare model risk-stratification. AUC was reported for individuals at risk up to the one-year mark (i.e., not censored and no event); five patients in our external test data met this criterion and were excluded from AUC calculation.

We found that survival prediction performance with a small dataset was erratic across timepoints, though performance generally increased with training data availability (Figure 2d). Generally, BrainIAC outperformed other models, with the exception of 20% data availability. At 10% training data availability (n=55), BrainIAC maintained high performance with an AUC of 0.67 (95% CI: 0.57-0.76), significantly surpassing the Scratch (p=0.0009) and MedicalNet (p=0.005) with AUC of 0.47 (95% CI: 0.37-0.57) and 0.50 (95% CI: 0.43-0.58), respectively. At 100% data availability, BrainIAC had the highest performance as well (AUC 0.68), which was on par with MedicalNet (0.65 and surpassed the Scratch model (0.52, p=0.004) (Figure 5h).

External testing showed similar performance trends, with BrainIAC outperforming MedicalNet and Scratch with stable performance at all data availability percentage, except 20%. At 10% training data BrainIAC resulted In AUC of 0.64 (95% CI: 0.55-0.73) significantly outperforming MedicalNet 0.55 (95% CI: 0.49-0.60) and Scratch 0.52 (95% CI: 0.41-0.62) (p=0.01). BrainIAC performance remained constant at 100% availability with AUC 0.62 improving over MedicalNet 0.59 and Scratch 0.51 (Figure 5i). The concordance index with 95% CI is reported in Supplementary Data Table 13-14 (Supplementary Data Table 15,16).

With 100% of training data available, BrainIAC and MedicalNet median risks scores were able to stratify patients into high-and low-risk groups, while the Scratch model was not (Figure 3). BrainIAC median risk scores significantly stratified high-risk and low-risk groups at all fine-tuning data availability thresholds (p<0.05 for each, Figure 3) on internal test set. In contrast, the Scratch model failed to achieve significant stratification at any threshold including 100% data availability. MedicalNet significantly stratified subjects at >=80% data availability, but failed at lower levels of data availability (Figure 3). On external testing, with 100% data BrainIAC and MedicalNet were able to achieve significant stratification, whereas Scratch model failed. BrainIAC was further able to perform significant stratification at 60% data availability compared to MedicalNet and Scratch.

**Figure 3.**
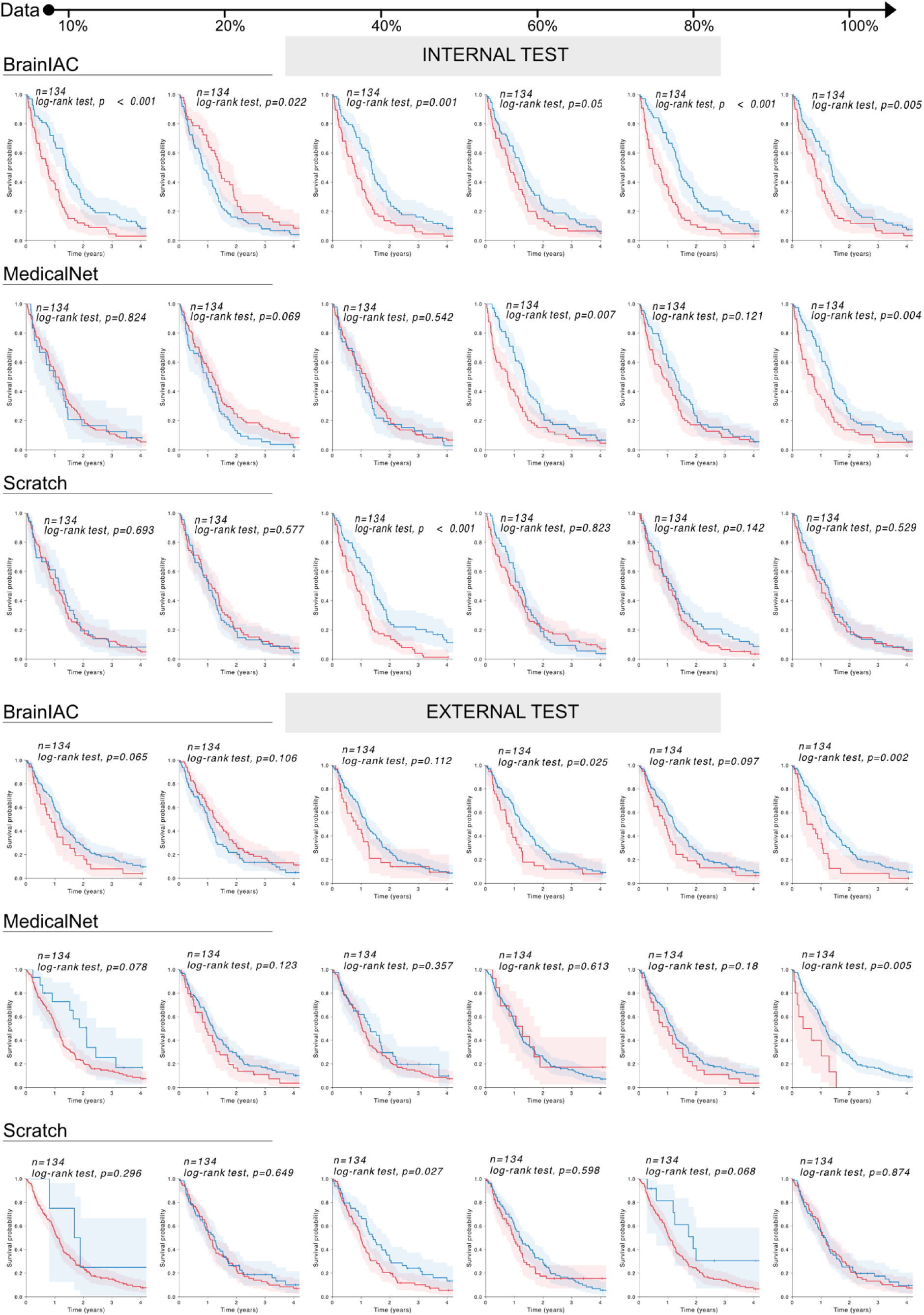
Kaplan Meier Curves for Overall Survival. For all three approaches (BrainIAC, MedicalNet and Scratch) across all data fractions, we performed risk stratification and generated Kaplan Meier curves on both internal (n=134) and external test (n=134) test sets. The stratification significance was calculated using the Log-rank test.

### Model Stability Analysis

To determine the stability and resiliency of different models to imaging artifacts and common perturbations seen across MRI scanners, we evaluated the performance of BrainIAC, MedicalNet and Scratch models across four downstream applications (MRI sequence classification, brain age prediction, tumor IDH mutation prediction, and survival in GBM) under varying levels of three imaging parameters: image contrast (scale: 0.5–2.0), Gibbs artifact (scale: 0.0–0.4), and bias field (scale: 0.0–0.4) at 100% data availability for fine-tuning.

BrainIAC generally demonstrated higher, more stable performance across perturbations of all three imaging parameters, when compared to MedicalNet and Scratch models (Figure 4). This most distinctly observed in tasks such as mutation prediction (n=396) and overall survival prediction (n=545) where the data availability is limited, and BrainIAC showed stable performance across perturbation scales whereas the MedicalNet and Scratch degraded significantly. For applications with ample data availability Sequence classification (n=5005) and brain age prediction (n=3882) the BrainIAC performance improvement is marginal and followed similar degradation trends compared to MedicalNet and Scratch.

**Figure 4.**
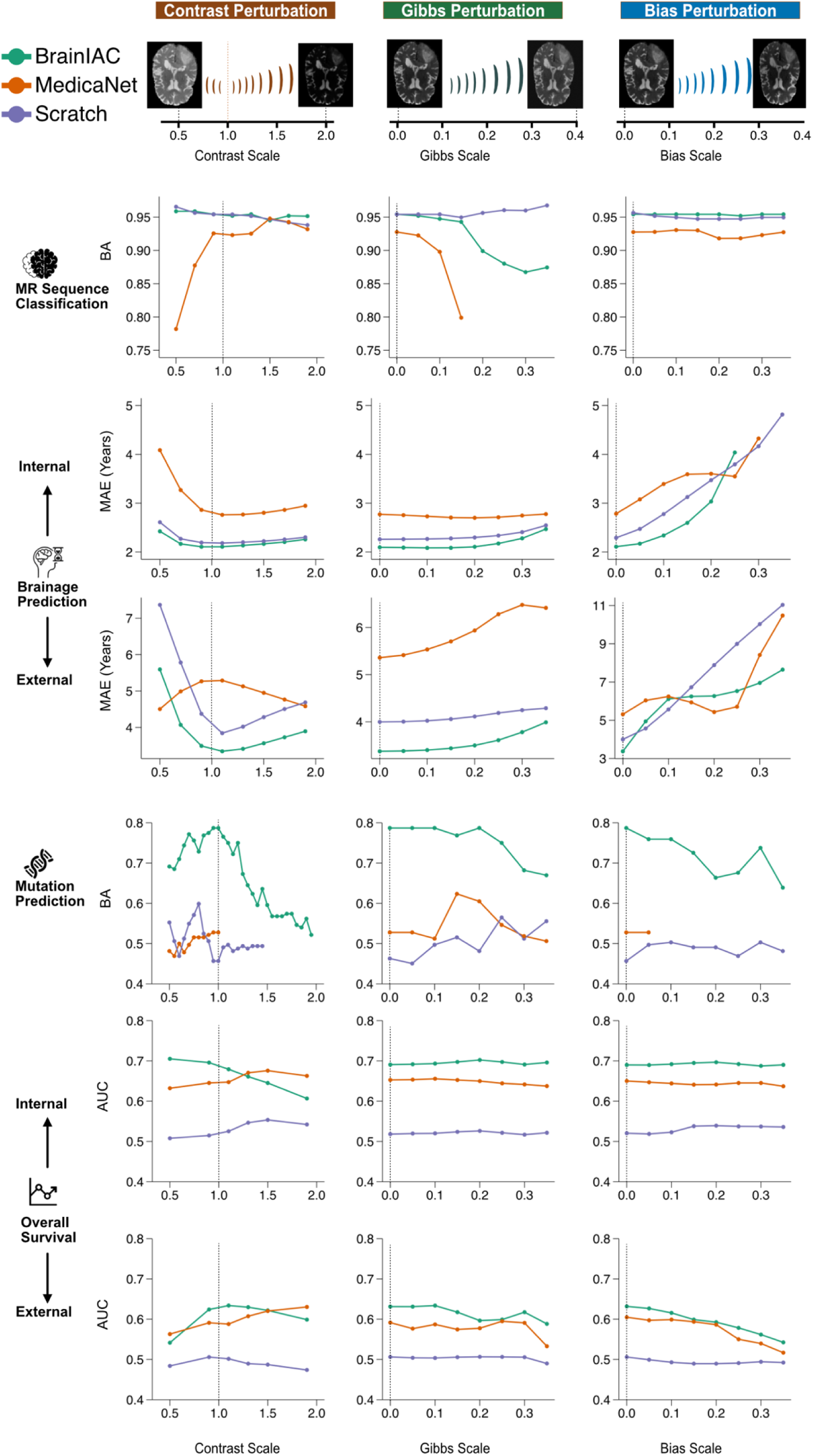
Perturbance performance comparison. Models trained on complete datasets were evaluated for stability against three types of imaging artifacts and perturbations (contrast, Gibbs, and bias) across all downstream applications. Performance was assessed across contrast scale variations (0.5-2.0, baseline=1.0), Gibbs artifacts (scale 0.0-0.4, baseline=0.0), and bias field perturbations (scale 0.0-0.4, baseline=0.0). The vertical dotted lines indicate baseline (no perturbation) performance for reference

Notably, each clinical task exhibited distinct vulnerability patterns: mutation prediction accuracy was particularly sensitive to contrast perturbations, while brain age prediction showed greater deterioration under bias perturbations. Sequence classification and overall survival prediction demonstrated greater resiliency, maintaining stable performance across all perturbation types.

## DISCUSSION

In this study, we present BrainIAC, a foundation model for brain MRI analysis developed with self-supervised contrastive pretraining and rigorously evaluated on 48,519 multiparametric brain MRI scans spanning multiple demographic and clinical settings. We find that BrainIAC consistently outperforms traditional supervised models and transfer learning from more general biomedical imaging models across multiple downstream applications on healthy and disease-containing scans with minimal fine-tuning. BrainIAC is robust to imaging perturbations. Our findings demonstrate BrainIAC’s adaptive and generalization capabilities, positioning it as a powerful foundation for development of clinically usable imaging-based deep learning tools, particularly in limited data scenarios.

The emergence of foundation models has advanced medical imaging artificial intelligence applications, from biomarker discovery to cancer diagnostics^10,39^. Several pretrained model frameworks exist for biomedical imaging^40,41^, but none are focused exclusively on Brain MRI, and only one, MedicalNet^27^ – which was used as a primary benchmark in this study – is able to analyze 3D images. While MedicalNet represents a powerful advance for biomedical imaging analysis, we find that a foundation model pretrained with SSL for multiparametric MRI consistently outperforms the broader biomedical imaging model. We hypothesize that the inherent differences between MRI intensity values, sequence acquisitions, and anatomy make a Brain MRI-specific foundation model critical to high-performing algorithms in neuroimaging. A few approaches to foundation models for Brain MRI lesion segmentation proposed^19,25^ and anomaly detection^42^, but the present work is the first to apply and rigorously evaluate an SSL approach for broad, classification problems, which represent important use cases for medical management of diseases that affect the brain.

BrainIAC was intentionally tested on tasks that have a range of perceived difficulty from a clinical standpoint. On one end of the spectrum, MRI sequence classification is straightforward for trained clinicians, and on the other end of the spectrum genomic subtyping and survival prediction are very challenging based on imaging alone. Supervised deep learning models have shown promise in even challenging brain MRI tasks^43,44^, but require a large amount of training data and are prone to performance degradation when applied in contexts outside that of which they were trained^45,46^, thus limiting their clinical usability and utility. BrainIAC showed consistently improved performance over other approaches in all tasks, regardless of perceived difficulty, particularly in low-data settings (<10% of data available). Notably, even with all training data made available, BrainIAC continued to demonstrate higher performance in tasks that were both challenging and had limited training cases available (i.e. mutational subtype and survival prediction), while in “easier” tasks with more data (i.e. sequence classification) the performance gap was narrower between BrainIAC and other approaches. This suggests the utility of BrainIAC as a foundation model is likely potentiated in settings of challenging tasks and low data, such as classification of rare cancers. Additionally, BrainIAC was found to be more generalizable for brain age and survival prediction, for which true external test sets were available for evaluation. For brain age prediction at 100% training data availability (3,882 scans), BrainIAC continued to show improved accuracy versus other approaches, suggesting that BrainIAC learned more informative, generalizable features to form a basis for fine-tuning. Hence, potentially serving as a feature extractor for imaging-based analysis of neurodegenerative and neurofunctional diseases. In tumor-related tasks, where labeled data scarcity often poses significant challenges, the improvement in performance provided by BrainIAC was clear.

Our study has several limitations. Our training and evaluations focused on four commonly used MRI sequences (T1-weighted, T2-weighted, T1CE, and FLAIR), but inclusion of other important sequence acquisition phases such as diffusion-weighted, dynamic contrast-enhancing, and fat suppression, may prove to be helpful in certain clinical contexts. The model was trained on skull-stripped images, thus limiting its application to intracranial analysis. We employed widely validated methods for both our neural network architecture (ResNet) and contrastive SSL pretraining (SimCLR). There may be room for further improvement with the use of newer SSL frameworks, such as DINO^3^ and MAE^4^, as well as alternative architectures such as vision transformers. While vision transformers have found success in 2D applications, such as digital pathology, 3D imaging analysis is often prohibitively computationally burdensome, though future work should explore these approaches. Finally, while this represents the largest pretrained brain MRI foundational model to-date, inclusion of even further training data may yield performance improvements. Future work will focus on investigation of performance improvements with incorporation of new training data, learning strategies and architectures.

In conclusion, BrainIAC represents a step towards generalized brain MRI analysis with self-supervised foundation models. With minimal fine-tuning, BrainIAC can raise the bar for performance on multiple MRI tasks. Our findings suggest that a BrainIAC foundation pipeline could replace traditional supervised learning strategies for brain MRI and allow for the development of models adaptable to challenging tasks in data-limited scenarios that were previously thought infeasible.

## METHODS

### Dataset

This study was conducted in accordance with the Declaration of Helsinki guidelines and following the approval of local Review Board (IRB). Waiver of consent was obtained from the IRB prior to research initiation due to use of public datasets and retrospective nature of the study. We curated a dataset pool of 48,519 brain MRI scans, including 24,504 T1W, 5389 T2W, 15372 T2FLAIR, and 3254 T1CE sequences. The data pool was aggregated from the following datasets : ABCD^47^ , ADNI , DFCI/BCH LGG, OASIS-3^48^, MCSA^49^, SOOP^50^, ABIDE^51^, CBTN LGG^52^, MIRIAD^53^, PPMI^54^, DLBS^55^, RadArt^56^, OASIS-2^57^, DFCI/BCH HGG, QIN-GBM^58^, RIDER^59^, UPENN-GBM^38^, BraTS 2023^60–63^, UCSF-PDGM^64^, wu1200^65^, LONG579^66^, BABY^67^, AOMIC^68^, Calgary^69^, HaN^70^, NIMH^71^, ICBM, IXI^72^, PING^73^, Pixar^74^, SALD^75^, PETfrog^76^ further details and acknowledgements are in Supplementary Data A.1

### Data preprocessing

We developed a systematic preprocessing pipeline to ensure standardization and quality control of the structural magnetic resonance imaging (MRI) data. Raw DICOM images were initially converted to NIFTI format using the dcm2nii package (Python v3.8). To address low-frequency intensity non-uniformity inherent in MRI acquisitions, we applied N4 bias field correction using SimpleITK. All scans were subsequently resampled to isotropic 1×1×1 mm³ voxels through linear interpolation, followed by rigid registration to the MNI space brain atlas. Finally, brain extraction/skull stripping was performed using the HD-BET package^77^ as the last preprocessing step before the analyses.

### Pretraining

We implemented a self-supervised pretraining approach based on SimCLR^5^, which has demonstrated great success in 3D radiological imaging analysis¹², for the training of the foundation model. We modified a ResNet50 architecture, from MONAI implementation, by removing the final classification layer and augmenting the backbone convolutional network with a projection head (multi-layer perceptron) post global average pooling layer to generate 2048-dimensional latent feature representations.

The contrastive learning framework employed a normalized temperature-scaled cross entropy (NT-Xent)^78^ loss function to optimize spatial learning. This approach maximized similarity between positive pairs (augmented views derived from the same image) while minimizing similarity between negative pairs (views from different images). Input volumes were standardized to (128,128,128) voxels at (1,1,1) mm spacing. We used a comprehensive augmentation pipeline comprising random flips, Gaussian blur, Gaussian noise, affine transformations (scale, rotation, translation), and random cropping, with subsequent resizing to maintain dimensional consistency (Figure 1b).

Model pretraining was conducted over 200 epochs with a batch size of 32 on an NVIDIA A6000 GPU, requiring approximately 72 hours for completion (Supplementary Data Table 17). Complete implementation details and code are available at [GitHub repository URL].

### Downstream Adaptation

We finetuned the foundation model for downstream adaptation across four distinct clinical applications: sequence classification, brain age prediction, mutation prediction, and overall survival analysis. We systematically evaluated model performance using three initialization strategies: Brain MRI foundation model (BrainIAC) fine-tuning, supervised training with random initialization (Scratch), and fine-tuning from MedicalNet weights (MedicalNet). Each application pipeline was constructed upon the vision encoder (ResNet50), with architectural modifications based on the specific task requirements (classification versus regression) and input characteristics (single versus multiple images).

For evaluation and comparison, we implemented a framework where the datasets were initially partitioned into training-validation and test-holdout sets. The training-validation data was further subdivided into multiple fractions (10%, 20%, 40%, 60%, 80%, and 100%) to assess model performance across varying data availability scenarios. For each fraction, we trained models using all three initialization approaches and evaluated their performance on the constant holdout set (Extended Data Figure 1). The architectural pipeline maintained a ResNet50 backbone, with task-specific modules appended for each application. The task-specific modules were randomly initialized across all approaches, and the ResNet50 weights were initialized according to the respective strategy: BrainIAC, MedicalNet, and Scratch. We report our results in accordance with the TRIPOD+AI statement guidance^79^.

### Sequence classification adaptation

For the sequence classification application, we formulated a four-way classification task to distinguish between T1, T2, FLAIR, and T1CE MR imaging sequences. The architecture comprises a ResNet50 vision encoder generating 2048-dimensional latent features, followed by a fully connected layer of matching dimensionality, followed by a four-neuron output layer. Training for all three approaches was implemented using cross-entropy loss under supervised conditions.

The model was optimized using Adam optimizer with approach-specific learning rates: 0.001 for scratch initialization and 0.0001 for both BrainIAC and MedicalNet approaches. Training was performed for 100 epochs with a batch size of 16, with a CrossEntropyLoss (Equation 1) and a Reduce-LR-On-Plateau learning rate scheduler. Model selection was based on validation set performance, with the best-performing checkpoints from each strategy retained for holdout set evaluation.

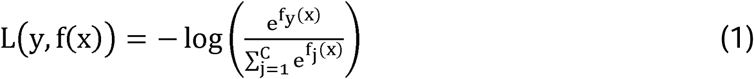

We utilized the BraTS23 dataset, comprising 5,880 images equally distributed across the four sequence classes. We quantified model performance using balanced accuracy (BA) as the primary metric. To evaluate the subgroup performance, we performed pairwise comparisons between sequences across all initialization strategies, using BA to quantify performance differences (Extended Data Figure 2).

### Brain age prediction adaptation

For the Brain age application, we formulated a regression task with Brain age being the regression variable. The pipeline consists of single input T1 image passed into a ResNet50 vision encoder followed by a fully connected layer of 2048 dimensions, which is further connected to a single neuron output for regression. The training is done in a supervised fashion with mean absolute error (MAE) as the primary loss function employed. We also use MAE as the performance evaluation metric. The individual data fraction models were trained using a batch size 16, with initial learning rate of 0.001(scratch) and 0.0001 (BrainIAC and MedicalNet) with a MSE loss (Equation 2), Adam optimizer and ReduceLROnPlateau learning rate scheduler for 100 epochs, and mean absolute error (MAE) as the evaluation metric.

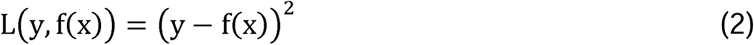

We aggregated a dataset pool of 14 datasets where we separated the dataset pool into a split of in-distribution (development) and out of distribution set. The in-distribution dataset consisted of (10) and out of distribution dataset (1072 images) consisted of (4). The in-distribution dataset was further pooled and split into a training-validation (3882 images) and holdout-testing split (1296 images) (with 80:20 ratio), where the training-validation split was further fragmented into dataset% fractions (Extended Data Figure 1). All the dataset fraction models were tested on in-distribution and out-of-distribution test sets.

### Mutation classification adaption

For the mutation classification application, we formulated the problem as a binary classification task for IDH mutation (IDH mutated Vs Wildtype) for diffuse glioma subjects from the UCSF-PDGM dataset containing 392 IDH mutated and 103 wildtype subjects. The pipeline, taking in two images (T1CE, FLAIR) from same subject as an input, consisted of the ResNet50 vision encoder which outputed two feature vectors (2048 dimensions) corresponding to the two input images. This was followed by the average pooling layers which results in a single feature vector of dimension 2048 that is passed into the fully connected layer with 2048 neurons and finally the output layer consisted of a single neuron for the binary classification tasks. The training procedure consisted of binary cross entropy loss (BCE) as the optimization metric along with Adam optimizer with the learning rate of 0.001 (Scratch) and 0.0001 (MedicalNet and BrainIAC) along with ReduceLROnPlateau scheduler. We used AUC as the performance evaluation and comparison metric along with balanced accuracy.

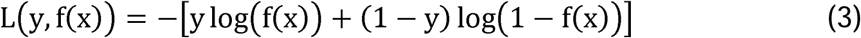

### Overall survival prediction

For overall survival prediction, we developed a classification pipeline to predict one-year overall survival outcomes in glioblastoma patients using the UPENN-GBM dataset, with 668 subjects— 360 of whom survived beyond one year. We reserved 534 subjects for training-validation and 134 as a holdout test set. We curated another external test set from TCGA-GBM/BraTS23 dataset of 134 subjects with complete survival information. We used 1-year overall survival as the primary endpoint. The pipeline processed four MRI modalities per subject: T1CE, FLAIR, T1, and T2 images. Each image is input into a ResNet50 vision encoder, generating four separate 2048-dimensional feature vectors which were pooled using average pooling layer generating a single 2048-dimensional feature vector. This unified vector was then passed through a fully connected layer with 2048 neurons, followed by an output layer of a single neuron for classification.

We computed a risk score for each patient based on the model’s output, using the median risk score from the training set as the threshold to stratify patients into high-risk and low-risk groups. Kaplan-Meier survival analysis and log-rank tests were conducted to evaluate the effectiveness of this stratification.

The pipeline was trained in a supervised fashion using a BCE loss and Adam optimizer. Learning rates were set at 0.001 (Scratch) and 0.0001 (MedicalNet and BrainIAC), along with a ReduceLROnPlateau scheduler over 200 epochs. The model performance was evaluated using AUC and Concordance Index (CI) metric. AUC was reported for individuals at risk up to the one-year mark (i.e., not censored and no event); five patients in our external test data met this criterion and were excluded from AUC calculation. Details about censoring can be found in Supplementary Data Table 12.

### Perturbation and Stability analysis

To simulate the real-world MRI acquisition and demographic differences, and evaluate the generalization capabilities of the finetuned models for the three approaches, we conducted perturbation and stability analyses. We selected three types of perturbations: contrast perturbation, Gibbs perturbation, and bias perturbation. For each downstream task, we chose the best-performing model (trained on 100% of the data) from the three approaches. During inference, each image in the test set was systematically perturbed across the entire range of each perturbation scale. After completing the inference sweep on the entire dataset, we then computed the model’s output using the corresponding task-specific metric. This procedure was repeated for all three perturbations, for each of the three best-performing models, across all four downstream tasks.

The contrast perturbation was applied using the AdjustContrast function from the MONAI framework, with a gamma scale ranging from -0.5 to 2, where 1 represents the normal condition. Gibbs perturbation was introduced using the GibbsNoise function from MONAI, with an alpha scale ranging from 0.0 to 0.4, and 0.0 representing the normal condition. Bias perturbation was performed using the RandBiasField function from MONAI, with a bias scale ranging from 0.0 to 0.4, where 0.0 denotes the normal state.

### Saliency Map and dimensionality reduction

To identify the regions in the images where the models focus most—leading to activation of model weights—we selected random images from the holdout set of each downstream application. We generated saliency maps for both the foundation model (BrainIAC) (Extended Data Figure 3) and the fine-tuned BrainIAC models adapted for each downstream task (Figure 5). We employed the smooth guided backpropagation approach to produce the saliency heatmaps, which involved stopping the negative gradients and allowing only activated neurons during backpropagation. This method is implemented in the MONAI framework as GuidedBackpropSmoothGrad.

**Figure 5.**
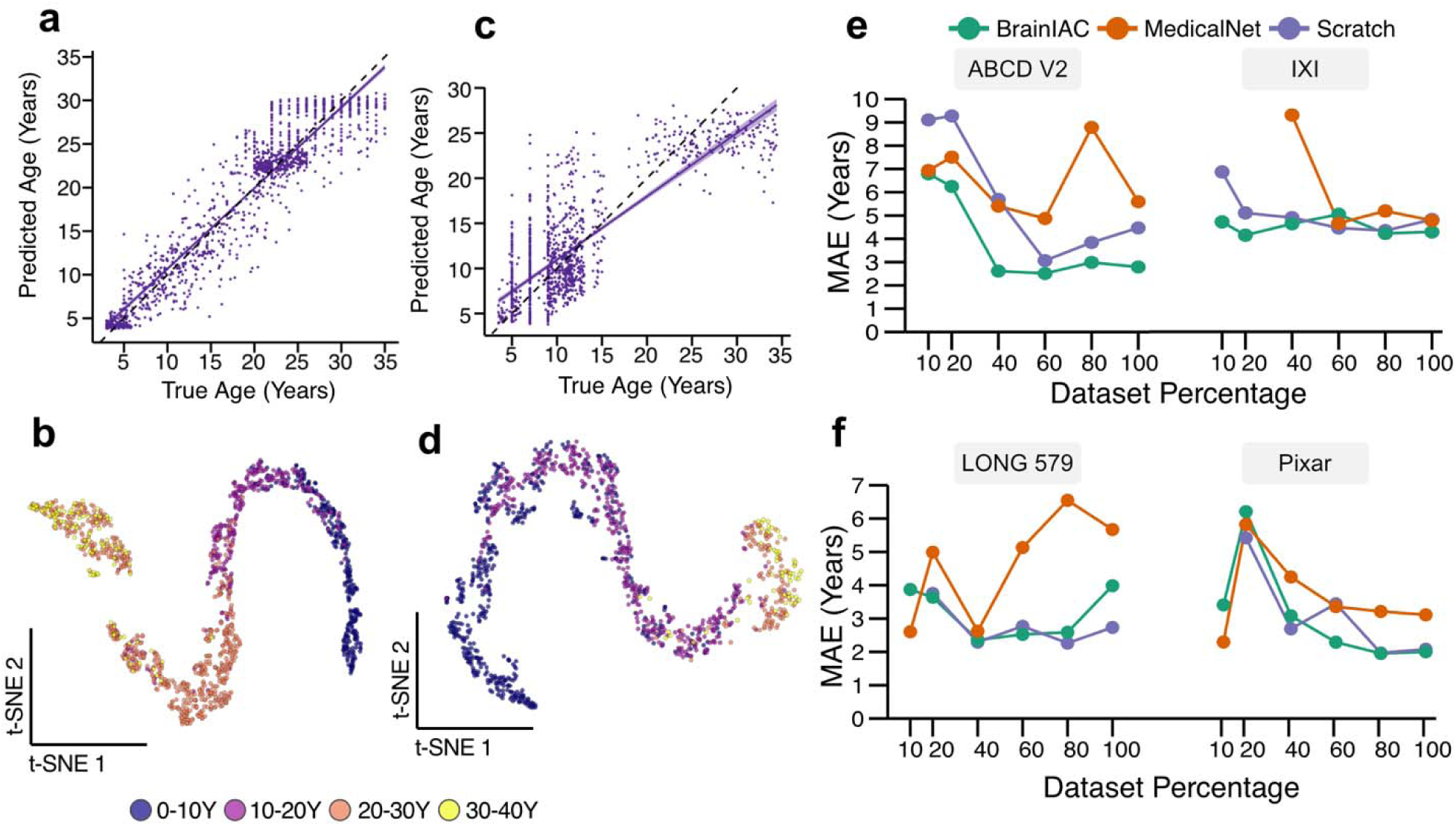
BrainIAC BrainAge prediction performance. **A** The scatter plot with regression line overlay represents the correlation between predicted age and true age for BrainIAC finetuned model (100%) for the internal test set (n=1296) **C**. and external test set (n=1072). The normal is denoted by a black dashed line. The t-SNE plots depicts the latent representation of the BrainIAC on the internal (**B**) and external (**D**) test sets. KNN clustering was performed K=4 bins (0-10 years, 10-20 years, 20-30 years, 30-40years) with Davies-Boulding index score of 0.475 for in-distribution test and 0.575 for external test. (**E, F**). The line plots shows the subgrpup performance comparison (MAE) on individual datasets from the external test set pool.

**Figure 6.**
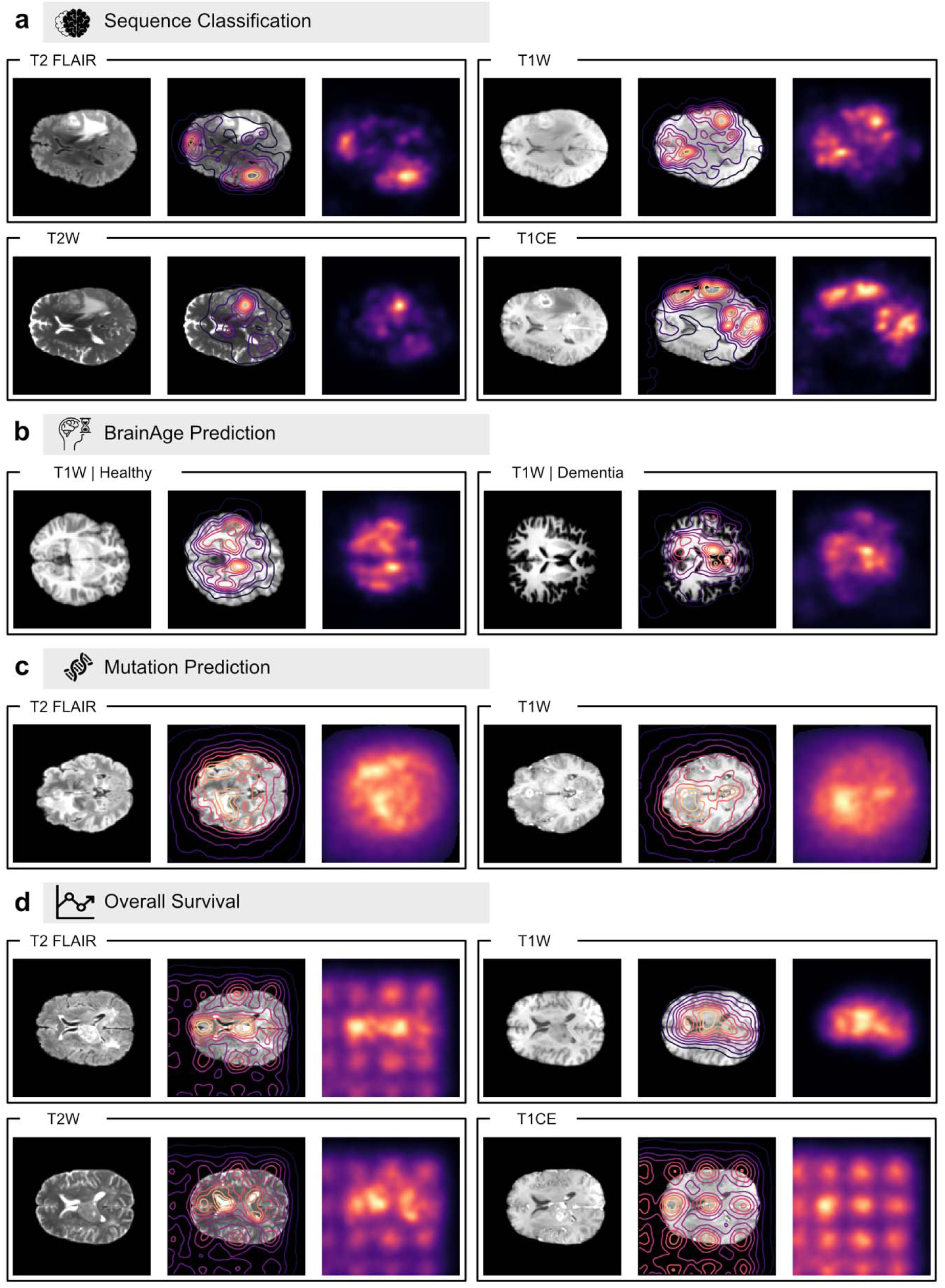
Saliency maps for BrainIAC models for downstream applications. Saliency maps showing model attention patterns across different clinical applications, with each panel displaying the original image (left), saliency map contour overlay (middle), and saliency heatmap (right). Saliency maps were generated for the BrainIAC finetuned model (100% data fraction) using smooth-guided backpropagation. **A** saliency map depiction for a single subject with T1CE, FLAIR, T1, and T2 sequences from the holdout test set for sequence classification application. **B** saliency map depiction on T1 sequence for two subjects from out-of-distribution set for brainage application. **C** saliency map depiction on T1CE and FLAIR sequences from a single subject from the holdout set for IDH mutation classification application. **D** saliency map depiction on T1CE, FLAIR, T1, and T2 sequences from a single subject from the holdout set for overall survival application.

We performed dimensionality reduction using t-distributed stochastic neighbor embedding (t-SNE) to visually represent the high-dimensional latent features (2048 dimensions). The features were reduced to two dimensions, and each point on the t-SNE map was color-coded based on age group binning for the Brain age task (Figure 4a), and sequence category for the sequence classification task (Figure 2b).

## List of abbreviations

BrainIAC: Brain Imaging Adaptive Core
CNN: Convolutional neural network
AUC: Area under the curve
BA: Balanced Accuracy
DL: Deep learning
MRI: Magnetic resonance imaging
SSL: Self-supervised Learning
HGG: High-grade glioma
LGG: Low-grade glioma
AI: Artificial Intelligence

## DATA AVAILABILITY

All data supporting the findings described in this manuscript are available in the article, in the Supplementary Information, and from the corresponding author upon request. The DFCI/BCH and RadART brain tumor dataset contains private hospital data that is controlled due to privacy concerns. Access to the derived dataset will be considered upon request to the corresponding author (Benjamin H. Kann, M.D., timeframe for response 2 weeks).

## CODE AVAILABILITY

The BrainIAC model is available on hugging face https://huggingface.co/Divytak/brainiac , the source code and downstream model weights will be made available at time of publication via Github repository.

## ACKNOWLEDGEMENTS

This study was supported in part by the National Institute of Health/ the National Cancer Institute (NIH/NCI) (U54 CA274516 and P50 CA165962), and Botha-Chan Low Grade Glioma Consortium. We would also like to acknowledge the Children’s Brain Tumor Network (CBTN) for the imaging and clinical data access, and ASCO Conquer Cancer Foundation: 2022A013157 Radiation Oncology Institute: ROI2022-9151

## EXTENDED DATA

**Extended Data Figure 1.**
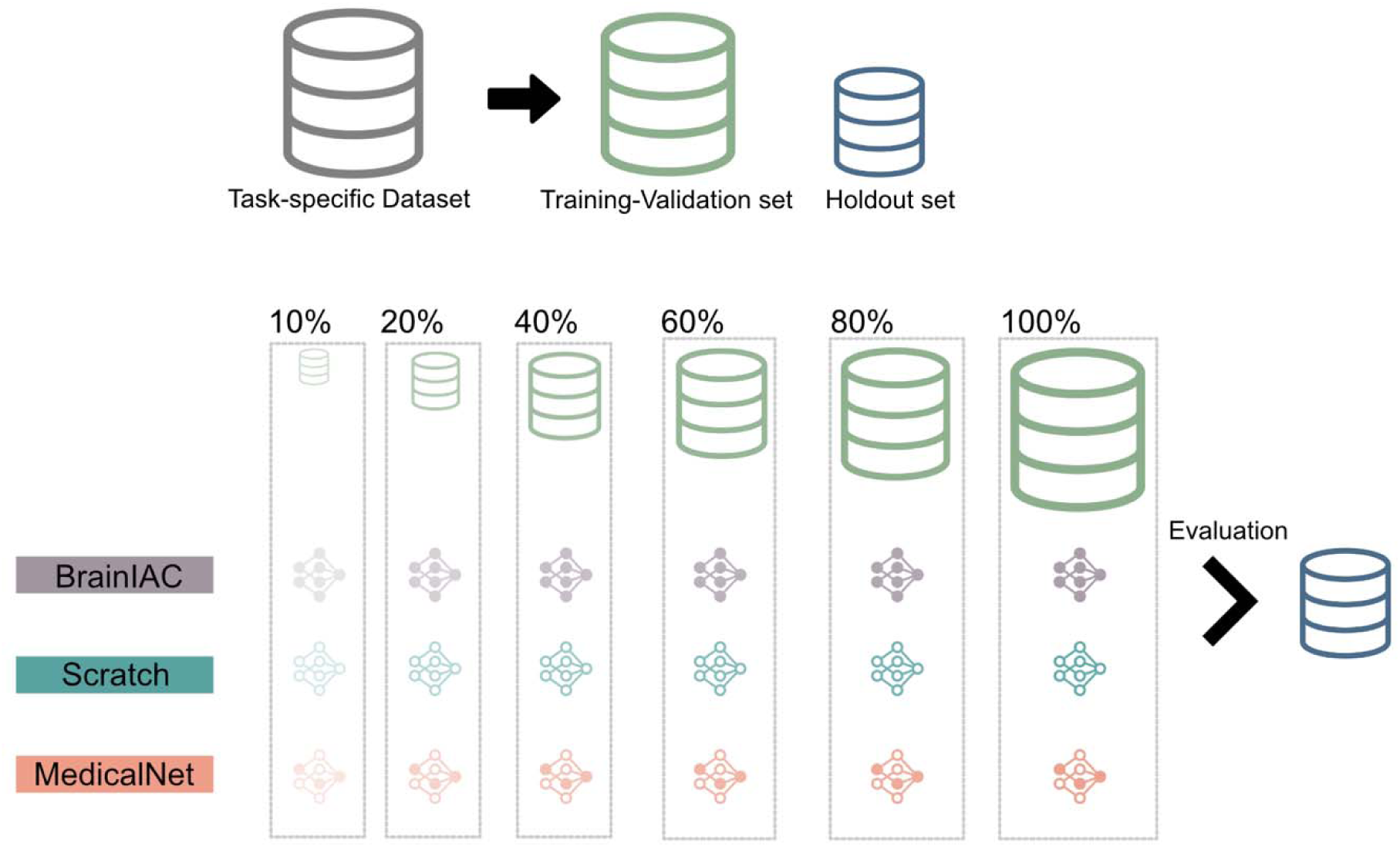
Downstream application adaption method. Each Downstream application is implemented by segregating the task specific dataset into a training-validation and test (holdout) sets, using the 80:20 split ratio. The training-validation set is further sampled to generate datasets of varying sample size (10% up to 100%). All three approaches are trained, separately, on each dataset faction and the resulting models are evaluated and compared on the reserved holdout set.

**Extended Data Figure 2.**
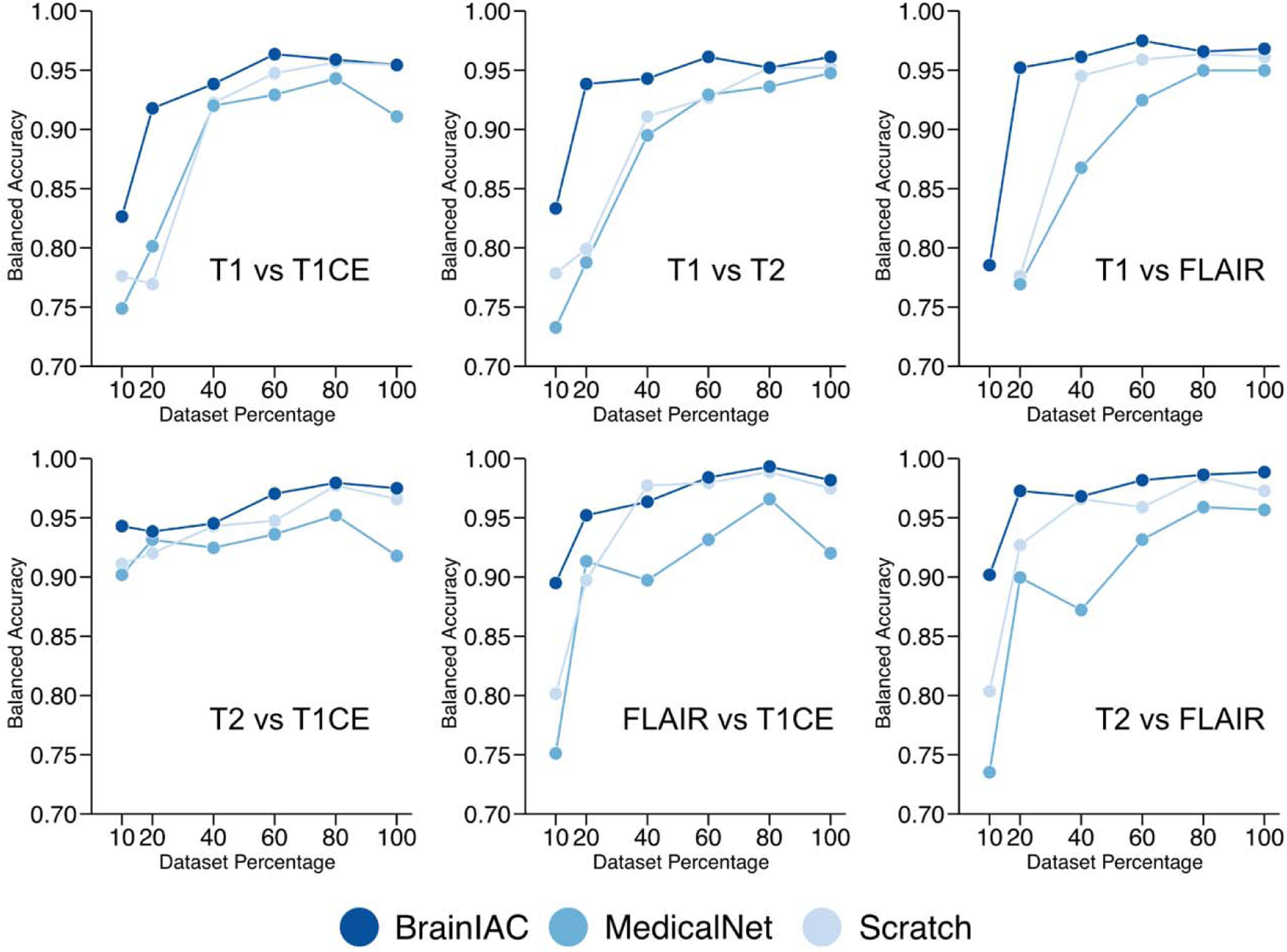
Sequence Classification Subgroup Analysis. The sequence classification application consisted of a 4-way classification between T1w, T2w. T1CE and FLAIR sequences. We further conduct a subgroup analysis on pairwise classification performance on the test set across the three approaches. Balanced accuracy is use as the evaluation metric for this analysis.

**Extended Data Figure 3.**
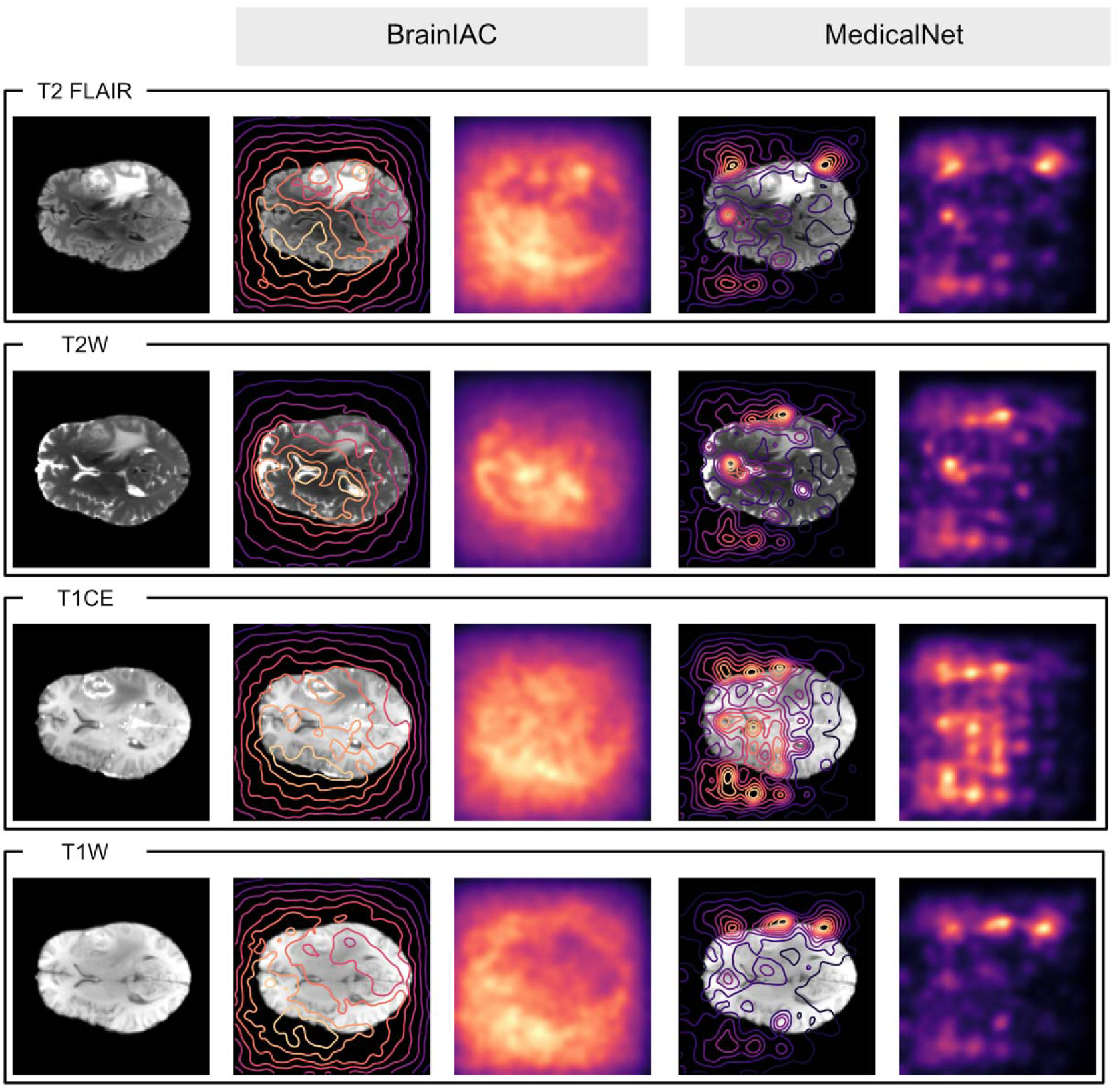
BrainIAC and MedicalNet saliency maps. We generated saliency maps and contour overlays for a ResNet50 model loaded with BrainIAC and MedicalNet weights. The saliency maps are generated for the 4 sequences of a random sample drawn from BraTS23 dataset.

