## Supplementary for "A foundation model for generalized brain MRI analysis"

| Dataset | Number of Images |
| --- | --- |
| ABCD | 7197 |
| ADNI | 4825 |
| DFCI/BCH LGG | 4245 |
| OASIS-3 | 4874 |
| MCSA | 2303 |
| SOOP | 3430 |
| ABIDE | 1099 |
| CBTN LGG | 1541 |
| MIRIAD | 523 |
| PPMI | 547 |
| DLBS | 670 |
| RadART LGG | 213 |
| OASIS-2 | 211 |
| DFCI/BCH HGG | 200 |
| QIN-GBM | 99 |
| RIDER | 38 |
| UPENN-GBM | 2520 |
| BRATS23 | 5880 |
| UCSF-PDGM | 990 |
| wu1200 | 1096 |
| LONG579 | 285 |
| BABY | 284 |
| AOMIC | 928 |
| Calgary | 345 |
| HAN | 318 |
| NIMH | 923 |
| ICBM | 809 |
| IXI | 155 |
| NYU | 152 |
| PING | 738 |
| Pixar | 132 |
| SALD | 181 |
| Petfrog | 268 |
| ABCDV2 | 500 |
| Total | 48,519 |

Supplementary Data Table 1. Dataset Image Distribution. In this study we aggregated 35 datasets comprising 48,519 Brain MRI scans from both public sources and private sources such as DFCI/BCH pediatric records, and the RadART study.

| Dataset | Link |
| --- | --- |
| ABCD | <https://nda.nih.gov/abcd> |
| ADNI | <https://adni.loni.usc.edu/data-samples/> |
| DFCI/BCH LGG | - |
| OASIS-3 | <https://sites.wustl.edu/oasisbrains/home/oasis-3/> |
| MCSA | <https://www.mayo.edu/research/clinical-trials/cls-20311806> |
| SOOP | <https://openneuro.org/datasets/ds004889/versions/1.1.2> |
| ABIDE | <https://fcon_1000.projects.nitrc.org/indi/abide/> |
| CBTN LGG | - |
| MIRIAD | <https://www.ucl.ac.uk/drc/research-clinical-trials/minimal-interval-resonance-imaging-alzheimers-disease-miriad> |
| PPMI | <https://www.ppmi-info.org/access-data-specimens/download-data> |
| DLBS | <https://fcon_1000.projects.nitrc.org/indi/retro/dlbs.html> |
| RadART LGG | - |
| OASIS-2 | <https://sites.wustl.edu/oasisbrains/home/oasis-2/> |
| DFCI/BCH HGG | - |
| QIN-GBM | <https://wiki.cancerimagingarchive.net/display/Public/QIN+GBM+Treatment+Response> |
| RIDER | <https://www.cancerimagingarchive.net/collection/rider-neuro-mri/> |
| UPENN-GBM | <https://www.cancerimagingarchive.net/collection/upenn-gbm/> |
| BRATS23 | <https://www.synapse.org/Synapse:syn51156910/wiki/621282> |
| UCSF-PDGM | <https://www.cancerimagingarchive.net/collection/ucsf-pdgm/> |
| wu1200 | <https://www.humanconnectome.org/study/hcp-young-adult/document/1200-subjects-data-release> |
| LONG579 | - |
| BABY | <https://www.sciencedirect.com/science/article/abs/pii/S1053811918302593?via%3Dihub> |
| AOMIC | <https://www.nature.com/articles/s41597-021-00870-6> |
| Calgary | <https://www.sciencedirect.com/science/article/pii/S2352340920301189> |
| HAN | - |
| HIMH | - |
| ICBM | <https://ida.loni.usc.edu/collaboration/access/appLicense.jsp> |
| IXI | <https://brain-development.org/ixi-dataset/> |
| NYU | - |
| PING | <https://pmc.ncbi.nlm.nih.gov/articles/PMC4628902/> |
| Pixar | - |
| SALD | <https://fcon_1000.projects.nitrc.org/indi/retro/sald.html#:~:text=The%20goals%20of%20the%20SALD,minute%20resting%20state%20fMRI%20scan> |
| Petfrog | - |
| ABCDV2 | <https://nda.nih.gov/abcd> |

Supplementary Data Table 2. Dataset Link. Summary and Information link for the publicly available datasets used in this study.

| Condition | Number of Images |
| --- | --- |
| healthy | 14,981 |
| Alzheimer's | 10,222 |
| PLGG | 5,999 |
| Dementia | 2,514 |
| Stroke | 3,430 |
| Autism | 1,099 |
| Parkinson's | 547 |
| HGG | 200 |
| GBM | 8,537 |
| Diffuse Glioma | 990 |

Supplementary Data Table 3. Dataset Disease Distribution. The aggregated dataset used for this study encompasses 10 medical settings from healthy to diffuse glioma.

| Dataset | Number of Images | T1w | T2w | T1CE | FLAIR |
| --- | --- | --- | --- | --- | --- |
| ABCD | 7197 | 7197 | - | - |  |
| ADNI | 4825 | 1844 | - | - | 2981 |
| DFCI/BCH LGG | 4245 | - | 1560 | - | 2685 |
| OASIS-3 | 4874 | 2283 | 1263 | - | 1328 |
| MCSA | 2303 | - | - | 522 | 1781 |
| SOOP | 3430 | 1715 | - | - | 1715 |
| ABIDE | 1099 | 1099 | - | - | - |
| CBTN LGG | 1541 | - | 466 | - | 1075 |
| MIRIAD | 523 | 523 | - | - | - |
| PPMI | 547 | 83 | - | - | 464 |
| DLBS | 670 | 335 | - | - | 335 |
| RadART | 213 | - | - | - | 213 |
| OASIS-2 | 211 | 211 | - | - | - |
| DFCI/BCH HGG | 200 | - | - | - | 200 |
| QIN-GBM | 99 | - | - | 99 | - |
| RIDER | 38 | - | - | 38 | - |

Supplementary Data Table 4. SSL Pretraining Dataset Distribution. BrainIAC was trained on 32,015 Images across 4 MR sequences (T1w, T2w, T1CE, FLAIR) pooled from 16 datasets.

| Category | Number of Images |
| --- | --- |
| T1 | 1470 |
| T2 | 1470 |
| T1CE | 1470 |
| FLAIR | 1470 |
| TOTAL | 5880 |

Supplementary Data Table 5. Dataset Sequence Distribution. The aggregated dataset used for this study encompassed 4 sequences T1w, T2w, T1CE, FLAIR.

| Data percentage | BrainIAC | MedicalNet | Scratch |
| --- | --- | --- | --- |
| 10 | 1.28 | 1.27 | 1.28 |
| 20 | 0.96 | 1.14 | 1.32 |
| 40 | 0.92 | 1.02 | 0.8 |
| 60 | 0.79 | 0.9 | 0.78 |
| 80 | 0.79 | 0.8 | 0.72 |
| 100 | 0.68 | 0.82 | 0.72 |

Supplementary Data Table 6. Davies-Bouldin Index clustering scores comparison for three approaches. KNN clustering (K=4) was performed on the latent features from ResNet50 (1024 dimensions) of the finetuned models, and Davies-Bouldin Index scores were calculated.

| Internal | External |
| --- | --- |
| SALD | ABCD V2 |
| Petfrog | Pixar |
| PING | Long579 |
| NYU | IXI |
| HAN | - |
| HIMH | - |
| Calgary | - |
| ICBM |  |
| AOMIC | - |
| WU1200 | - |

Supplementary Data Table 7. Brain age Dataset Split. Brain age dataset pool was split into internal (developmental) and external (out of distribution) sets. The internal set was further split into the training-validation and internal (holdout) test set with 80:20 split ratio each. The Internal and external test sets were used for model performance evaluation and comparison.

| Age Bin (years) | Number of Images |
| --- | --- |
| Internal Set | |
| 0-10 | 1178 |
| 10 20 | 1139 |
| 20-30 | 2224 |
| 30-40 | 636 |
| External Set | |
| 0-10 | 498 |
| 10 20 | 393 |
| 20-30 | 121 |
| 30-40 | 60 |

Supplementary Data Table 8. Age Distribution of Brain age Dataset. The Internal development set and the external test set consists of subjects with age ranging from 1-40, binned into 4 categories – 0-10Y, 10-20Y, 20-30Y, 30-40Y.

|  | UCSF-PDGM |
| --- | --- |
| Age : median(range) | 59 (17-94) |
| Sex – M n(%) | 295(59.5%) |
| Sex – F (%) | 200(40.5%) |
| Vital Status – Dead n(%) | 247(49%) |
| Vital Status – Alive n(%) | 248(51%) |
| IDH-wildtype n(%) | 392 (79.9%) |
| IDH-mutant n(%) | 103 (20.1%) |

Supplementary Data Table 9. UCSF-PDGM dataset patient characteristics for IDH mutation classification task.

| Data Percentage | Scratch | MedicalNet |
| --- | --- | --- |
| 10 | 0.11 | 0.18 |
| 20 | 0.04 | 0.34 |
| 40 | 0.072 | 0.16 |
| 60 | 0.002 | 0.823 |
| 80 | 0.0001 | 0.04 |
| 100 | 0.014 | 0.173 |

Supplementary Data Table 10. P values for IDH mutation classification AUC across three approaches on internal dataset. The P values were calculated in pair with AUC of BrainIAC using DeLong test.

| Percentage | BrainIAC | MedicalNet | Scratch |
| --- | --- | --- | --- |
| FLAIR | | | |
| 10 | 30.47 | 29.75 | 31.74 |
| 20 | 29.52 | 31.25 | 32 |
| 40 | 29.36 | 30.23 | 32.03 |
| 60 | 29.33 | 31 | 30.96 |
| 80 | 28.58 | 31.8 | 32.03 |
| 100 | 28.7 | 31.82 | 31.32 |
| T1CE | | | |
| 10 | 30.73 | 29.41 | 31.71 |
| 20 | 29.81 | 32.36 | 32 |
| 40 | 30.21 | 33.22 | 32.06 |
| 60 | 29.14 | 40.8 | 31.11 |
| 80 | 29.61 | 40.81 | 31.9 |
| 100 | 28.73 | 32.64 | 31.29 |

Supplementary Data Table 11. ComDIST comparison for IDH mutation prediction for BrainIAC, MedicalNet and Scratch on test set.

|  | | UPENN-GBM | Brats23/TCGA-GBM |
| --- | --- | --- | --- |
| Age: Median (Range) | | 63.0(18.7- 88.5) | 59.0(17.0-84.0) |
| Sex, No | |  |  |
|  | Male | 403 | 83 |
|  | Female | 265 | 51 |
| Survival Time in Days, Median,(Range) | | 384(3- 6160) | 383(5-2768) |
| Survival Status | |  |  |
|  | Deceased | 644 | 121 |
|  | Alive | 17 | 13 |
|  | Lost to Follow-up | 7 | - |

Supplementary Data Table 12. UPENN-GBM and Brats23/TCGA-GBM dataset patient characteristics.

| Concordance Index (CI) | Lower 95% CI | Upper 95% CI |
| --- | --- | --- |
| Scratch | | |
| 0.51 | 0.45 | 0.57 |
| 0.50 | 0.45 | 0.57 |
| 0.58 | 0.52 | 0.64 |
| 0.53 | 0.47 | 0.58 |
| 0.57 | 0.50 | 0.63 |
| 0.51 | 0.45 | 0.57 |
| BrainIAC | | |
| 0.58 | 0.52 | 0.64 |
| 0.45 | 0.39 | 0.50 |
| 0.60 | 0.54 | 0.66 |
| 0.56 | 0.50 | 0.62 |
| 0.58 | 0.53 | 0.64 |
| 0.57 | 0.51 | 0.63 |
| MedicalNet | | |
| 0.53 | 0.50 | 0.57 |
| 0.51 | 0.45 | 0.57 |
| 0.51 | 0.46 | 0.58 |
| 0.53 | 0.47 | 0.59 |
| 0.54 | 0.48 | 0.60 |
| 0.58 | 0.53 | 0.64 |

Supplementary Data Table 13. Concordance Index of 1-year overall survival for Brats23/TCGA-GBM dataset. The performance of the three approaches (BrainIAC, MedicalNet, and Scratch) are compared across the varying of data availability setting from 10% to 100%. 95% confidence intervals (CIs) were calculated using 1,000 bootstrap samples.

| Concordance Index (CI) | Lower 95% CI | Upper 95% CI |
| --- | --- | --- |
| Scratch | | |
| 0.47 | 0.42 | 0.53 |
| 0.47 | 0.41 | 0.53 |
| 0.60 | 0.55 | 0.66 |
| 0.53 | 0.48 | 0.59 |
| 0.55 | 0.48 | 0.61 |
| 0.54 | 0.48 | 0.60 |
| BrainIAC | | |
| 0.64 | 0.59 | 0.69 |
| 0.42 | 0.37 | 0.48 |
| 0.63 | 0.58 | 0.69 |
| 0.59 | 0.54 | 0.65 |
| 0.66 | 0.60 | 0.71 |
| 0.63 | 0.56 | 0.68 |
| MedicalNet | | |
| 0.50 | 0.46 | 0.55 |
| 0.43 | 0.38 | 0.49 |
| 0.47 | 0.41 | 0.53 |
| 0.59 | 0.54 | 0.64 |
| 0.56 | 0.51 | 0.60 |
| 0.60 | 0.55 | 0.65 |

Supplementary Data Table 14. Concordance Index of 1-year overall survival for UPENN-GBM dataset. The performance of the three approaches (BrainIAC, MedicalNet, and Scratch) are compared across the varying of data availability setting from 10% to 100%. 95% confidence intervals (CIs) were calculated using 1,000 bootstrap samples.

| Data percentage | MedicalNet | Scratch |
| --- | --- | --- |
| 10 | 0.0056 | 0.0009 |
| 20 | 0.51 | 0.24 |
| 40 | 0.0001 | 0.52 |
| 60 |  | 0.24 |
| 80 | 0.008 | 0.001 |
| 100 | 0.48 | 0.004 |

Supplementary Data Table 15. P-Values for AUC comparison across three approaches on internal test set. P values were calculated by comparing AUCs using the DeLong test.

| Data percentage | MedicalNet | Scratch |
| --- | --- | --- |
| 10 | 0.08 | 0.01 |
| 20 | 0.11 | 0.34 |
| 40 | 0.01 | 0.49 |
| 60 | 0.49 | 0.73 |
| 80 | 0.05 | 0.39 |
| 100 | 0.64 | 0.03 |

Supplementary Data Table 16. P-Values for AUC comparison across three approaches on external test set. P values were calculated by comparing AUCs using the DeLong test.

| Hyper-Parameter | Value |
| --- | --- |
| Model | |
| Convolutional layers | 50 |
| FC layers | 2 |
| Parameters (million) | 52.5 |
| Latent dimension | 1024 |
| Model size (mb) | 209.8 |
| Input shape (H,W,D) | 128,128,128 |
| Transforms | |
| Crop scale (min,max) | 0.2,1 |
| Resizing mode | trilinear |
| Rotate range (min, max) (radian) | -0.34 - 0.34 |
| Translate range (pixels) | -15 - 15 |
| Scale range | 0.0 – 1.0 |
| Flip axis | 0,1,2 |
| Gaussian smooth range (sigma) | 0.25 – 1.5 |
| Shift intensity range | 50 – 100 |
| Contrast range (gamma) | 0.5 – 2.0 |
| Gaussian smooth range (std) | 0.0 – 0.09 |
| Training | |
| Loss | NTXent |
| Scheduler | Cosine |
| Learning Rate | 0.0001 |
| Momentum | 0.9 |
| Decay | 0.00005 |
| Optimizer | SGD |

Supplementary Data Table 17. SimCLR pretraining hyperparameters. ResNet50 was chosen as the convolutional backbone, the pretraining was done on 1* 40GB Nvidia A6000 GPU.

**A.1 Primary Datasets**

**ABCD**

Data used in the preparation of this article were obtained from the Adolescent Brain Cognitive Development SM (ABCD) Study (https://abcdstudy.org), held in the NIMH Data Archive (NDA). This is a multisite, longitudinal study designed to recruit more than 10,000 children age 9-10 and follow them over 10 years into early adulthood. The ABCD Study® is supported by the National Institutes of Health and additional federal partners under award numbers U01DA041048, U01DA050989, U01DA051016, U01DA041022, U01DA051018, U01DA051037, U01DA050987, U01DA041174, U01DA041106, U01DA041117, U01DA041028, U01DA041134, U01DA050988, U01DA051039, U01DA041156, U01DA041025, U01DA041120, U01DA051038, U01DA041148, U01DA041093, U01DA041089, U24DA041123, U24DA041147. A full list of supporters is available at https://abcdstudy.org/federal-partners.html. A listing of participating sites and a complete listing of the study investigators can be found at https://abcdstudy.org/consortium_members/. ABCD consortium investigators designed and implemented the study and/or provided data but did not necessarily participate in the analysis or writing of this report. This manuscript reflects the authors' views and may not reflect the opinions or views of the NIH or ABCD consortium investigators. The ABCD data repository grows and changes over time. The ABCD data used in this report came from the fast-track data release. The raw data are available at https://nda.nih.gov/edit_collection.html?id=2573. Instructions on how to create an NDA study are available at https://nda.nih.gov/training/modules/study.html). Additional support for this work was made possible from supplements to U24DA041123 and U24DA041147, the National Science Foundation (NSF 2028680), and Children and Screens: Institute of Digital Media and Child Development Inc. (Casey et al., 2018)

**ADNI**

**DFCI/BCH LGG**

The DFCI/BCH brain tumor dataset contains private hospital data that is controlled due to privacy concerns. Access to the derived dataset will be considered upon request to the corresponding author (Benjamin H. Kann, M.D.,, timeframe for response 2 weeks).

**OASIS-3**

Data were provided by OASIS , OASIS-3: Longitudinal Multimodal Neuroimaging: Principal Investigators: T. Benzinger, D. Marcus, J. Morris; NIH P30 AG066444, P50 AG00561, P30 NS09857781, P01 AG026276, P01 AG003991, R01 AG043434, UL1 TR000448, R01 EB009352. AV-45 doses were provided by Avid Radiopharmaceuticals, a wholly owned subsidiary of Eli Lilly. OASIS-3_AV1451: Principal Investigators: T. Benzinger, J. Morris; NIH P30 AG066444, AW00006993. AV-1451 doses were provided by Avid Radiopharmaceuticals, a wholly owned subsidiary of Eli Lilly.

**MCSA**

The data contained in this analysis were obtained under research grant U01 AG006786 (PI: Ronald C. Petersen, M.D., Ph. D.) from the National Institutes of Health to the Mayo Clinic Study of Aging

**ABIDE**

Primary support for the work by Adriana Di Martino was provided by the ([NIMH K23MH087770](https://projectreporter.nih.gov/project_info_details.cfm?aid=7772415)) and the Leon Levy Foundation.

Primary support for the work by Michael P. Milham and the INDI team was provided by gifts from Joseph P. Healy and the Stavros Niarchos Foundation to the Child Mind Institute, as well as by an NIMH award to MPM ( [NIMH R03MH096321](https://projectreporter.nih.gov/project_info_details.cfm?aid=8241553)).

**CBTN LGG**

The CBTN data is available upon request at “ <https://cbtn.org/> “

**MIRIAD**

Data used in the preparation of this article were obtained from the MIRIAD database. The MIRIAD investigators did not participate in analysis or writing of this report. The MIRIAD dataset is made available through the support of the [UK Alzheimer's Society](http://www.alzheimers.org.uk/) (Grant RF116). The original data collection was funded through an unrestricted educational grant from GlaxoSmithKline (Grant 6GKC)."

**PPMI**

Data used in the preparation of this article was obtained on [2024-08-15] from the Parkinson’s Progression Markers Initiative (PPMI) database (www.ppmi-info.org/access-dataspecimens/download-data), RRID:SCR_006431. For up-to-date information on the study, visit www.ppmi-info.org. PPMI – a public-private partnership – is funded by the Michael J. Fox Foundation for Parkinson’s Research and funding partners, including 4D Pharma, Abbvie, AcureX, Allergan, Amathus Therapeutics, Aligning Science Across Parkinson's, AskBio, Avid Radiopharmaceuticals, BIAL, BioArctic, Biogen, Biohaven, BioLegend, BlueRock Therapeutics, Bristol-Myers Squibb, Calico Labs, Capsida Biotherapeutics, Celgene, Cerevel Therapeutics, Coave Therapeutics, DaCapo Brainscience, Denali, Edmond J. Safra Foundation, Eli Lilly, Gain Therapeutics, GE HealthCare, Genentech, GSK, Golub Capital, Handl Therapeutics, Insitro, Jazz Pharmaceuticals, Johnson & Johnson Innovative Medicine, Lundbeck, Merck, Meso Scale Discovery, Mission Therapeutics, Neurocrine Biosciences, Neuron23, Neuropore, Pfizer, Piramal, Prevail Therapeutics, Roche, Sanofi, Servier, Sun Pharma Advanced Research Company, Takeda, Teva, UCB, Vanqua Bio, Verily, Voyager v. 25MAR2024 Therapeutics, the Weston Family Foundation and Yumanity Therapeutics.

**OASIS-2**

OASIS-2: Longitudinal: Principal Investigators: D. Marcus, R, Buckner, J. Csernansky, J. Morris; P50 AG05681, P01 AG03991, P01 AG026276, R01 AG021910, P20 MH071616, U24 RR021382

**DFCI/BCH HGG**

The DFCI/BCH brain tumor dataset contains private hospital data that is controlled due to privacy concerns. Access to the derived dataset will be considered upon request to the corresponding author (Benjamin H. Kann, M.D.,, timeframe for response 2 weeks).

**UPENN-GBM**

Bakas, S., Sako, C., Akbari, H., Bilello, M., Sotiras, A., Shukla, G., Rudie, J. D., Flores Santamaria, N., Fathi Kazerooni, A., Pati, S., Rathore, S., Mamourian, E., Ha, S. M., Parker, W., Doshi, J., Baid, U., Bergman, M., Binder, Z. A., Verma, R., … Davatzikos, C. (2021). Multi-parametric magnetic resonance imaging (mpMRI) scans for de novo Glioblastoma (GBM) patients from the University of Pennsylvania Health System (UPENN-GBM) (Version 2) [Data set]. The Cancer Imaging Archive. <https://doi.org/10.7937/TCIA.709X-DN49>

**Wu1200**

This HCP data release includes high-resolution 3T MR scans from young healthy adult twins and non-twin siblings (ages 22-35) using four imaging modalities: structural images (T1w and T2w), resting-state fMRI (rfMRI), task-fMRI (tfMRI), and high angular resolution diffusion imaging (dMRI). Behavioral and other individual subject measure data (both NIH Toolbox and non-Toolbox measures) is available on all subjects. MEG data and 7T MR data is available for a subset of subjects (twin pairs). The Open Access Dataset includes imaging data and most behavioral data. All details in the imaging protocols can be found at study webpage (<https://humanconnectome.org/study/hcpyoung-adult/document/1200-subjects-data-release/>)

**LONG579**

The public neuroimaging and behavioral dataset entitled “A longitudinal neuroimaging dataset on language processing in children ages 5, 7, and 9 years old” available on the OpenNeuro project (https://openneuro.org) and organized in compliance with the Brain Imaging Data Structure (BIDS). It includes 322 participants, recruited from the Austin, Texas. All neuroimaging data were collected using a Siemens Skyra 3 T MRI scanner located at The University of Texas at Austin Imaging Research Center. All images were acquired using a 64-channel head coil. Participants were positioned supine in the MRI scanner and foam pads were placed around the head to minimize movement. T1- weighted Magnetization Prepared - RApid Gradient Echo (MPRAGE) images were collected using GRAPPA, a parallel imaging technique based on k-space, and the following parameters: GRAPPA accel.factor PE = 2, TR = 1900 ms, TE = 2.43 ms, field of view = 256 mm, matrix size = 256 × 256, bandwidth = 180 Hz/Px, slice thickness = 1 mm, number of slices = 192, voxel size = 1 mm isotropic, flip angle = 9°. (Wang et al., 2022)

**BABY**

The Baby Connectome Project (BCP: https://nda.nih.gov/edit_collection.html?id=2848) is a four-year study of children from birth through five years of age, intended to provide a better understanding of how the brain develops from infancy through early childhood and the factors that contribute to healthy brain development. This project is a research initiative of the Neuroscience Blueprint – a cooperative effort among the 15 NIH Institutes, Centers, and Offices that support neuroscience research. The BCP is supported by Wyeth Nutrition through a donation to the FNIH. Images are acquired on 3T Siemens Prisma MRI scanners using a Siemens 32-channel head coil at the Center for Magnetic Resonance Research (CMRR) at the University of Minnesota and the Biomedical Research Imaging Center (BRIC) at the University of North Carolina at Chapel Hill (Howell et al., 2019).

**AOMIC**

The Amsterdam Open MRI Collection (AOMIC, https://openneuro.org/datasets/ds003097/versions/1.2.1) is a collection of three datasets with multimodal (3T) MRI data, including structural (T1-weighted), diffusionweighted, and (resting-state and task-based) functional BOLD MRI data, as well as detailed demographics and psychometric variables from a large set of healthy participants (N = 928, N = 226, and N = 216). Data from all three datasets were acquired on the same Philips 3T scanner (Philips, Best, the Netherlands) but underwent several upgrades in between the three studies (Snoek et al., 2021).

**Calgary**

The Preschool MRI study in The Developmental Neuroimaging Lab at the University of Calgary uses different magnetic resonance imaging (MRI) techniques to study brain structure and function in early childhood (https://osf.io/axz5r/files/osfstorage). All imaging for this dataset was conducted using the same General Electric 3T MR750w system and 32-channel head coil (GE, Waukesha, WI) at the Alberta Children’s Hospital in Calgary, Canada. Children were scanned either while awake and watching a movie, or while sleeping without sedation. The University of Cal- gary Conjoint Health Research Ethics Board (CHREB) approved this study (REB13-0020). T1-weighted images were acquired using an FSPGR BRAVO sequence with TR = 8.23 ms, TE = 3.76 ms, TI = 540 ms, flip angle=12 degrees, voxel size = 0.9x0.9x0.9 mm3, 210 slices, matrix size=512x512, field of view=23.0 cm. ASL images were acquired with the vendor supplied pseudo continuous 3D ASL sequence with TR = 4.56 s, TE = 10.7 ms, in-plane reso- lution of 3.5x3.5 mm2, post label delay of 1.5 s, and thirty 4.0 mm thick slices. The sequence scan time was 4.4 minutes (Reynolds et al., 2020)

**NIMH**

The data used in this work was collected from the 5.1 release (https://nda.nih.gov/edit_collection.html?id=1151) . MRI scans were acquired using either General Electric or Siemens 1.5 Tesla scanners involving six sites or Pediatric Study Centers (PSC) in the United States. The Institutional Review Board at the University of Wisconsin-Madison also approved the analysis of the data of this human subject. Sequence type: 3D FLASH/SPGR; GE sequence: pulse sequence=SPGR, mode=3D; TR: 22 ms; TE: 10-11 ms; excitation pulse angle: 30 degrees; orientation: sagittal; FoV: 250mmISx250mmAP; matrix: 256 x 256 ( x 124 - 180 slices); slices: 160- 180 slices of 1-1.5 mm thickness (cover entire head). Note that on GE systems with a 124-slice limitation, slice thickness should be adjusted to cover the entire head with 124 slices: signal averages: 1; scan time: 11.6 – 16.8 min (Evans, 2006).

**ICBM**

Data used in the preparation of this work were obtained from the International Consortium for Brain Mapping (ICBM) database (www.loni.usc.edu/ICBM). The ICBM project (Principal Investigator John Mazziotta, M.D., University of California, Los Angeles) is supported by the National Institute of Biomedical Imaging and BioEngineering. ICBM is the result of efforts of co-investigators from UCLA, Montreal Neurologic Institute, University of Texas at San Antonio, and the Institute of Medicine, Juelich/Heinrich Heine University - Germany. Data collection and sharing for this project was provided by the International Consortium for Brain Mapping (ICBM; Principal Investigator: John Mazziotta, MD, PhD). ICBM funding was provided by the National Institute of Biomedical Imaging and BioEngineering. ICBM data are disseminated by the Laboratory of Neuro Imaging at the University of Southern California (Kötter et al., 2001).

**IXI**

The data has been collected at three different hospitals in London:Hammersmith Hospital using a Philips 3T system (details of scanner parameters: http://braindevelopment.org/scanner-philips-medical-systems-intera-3t/), Guy’s Hospital using a Philips 1.5T system (details of scanner parameters: http://braindevelopment.org/scanner-philips-medical-systems-gyroscan-intera-1-5t/), Institute of Psychiatry using a GE 1.5T system (details of the scan parameters not available at the moment). The Thames Valley MREC granted ethical approval. The T1 and T2 images were acquired prior to diffusion-weighted imaging using 3D MRPRAGE and dual-echo weighted imaging (IXI Dataset – Brain Development, n.d.).

**NYU2(CoRR)**

The Consortium for Reliability and Reproducibility (CoRR, http://fcon_1000.projects.nitrc.org/fcpClassic/FcpTable.html ) has aggregated 1,629 typical individuals’ resting state fMRI (rfMRI) data (5,093 rfMRI scans) from 18 international sites and is openly sharing them via the International Data-sharing Neuroimaging Initiative (INDI). In this study, we used a subset from CoRR study ”NYU 2” created by New York University (Di Martino, Kelly)(An Open Science Resource for Establishing Reliability and Reproducibility in Functional Connectomics | Scientific Data, n.d.).

**PING**

The PING Data Resource(https://nda.nih.gov/edit_collection.html?id=2607) is the product of a multi-site project involving developmental researchers across the United States, including UC San Diego, the University of Hawaii UC Los Angeles Children’s Hospital of Los Angeles of the University of Southern California UC Davis Kennedy Krieger Institute of Johns Hopkins University Sackler Institute of Cornell University University of Massachusetts Massachusetts General Hospital at Harvard University and Yale University. The Data Resource includes neurodevelopmental histories, information about developing mental and emotional functions, multimodal brain imaging data, and genotypes for well over 1000 children and adolescents between the ages of 3 and 20. The PING imaging protocol takes advantage of key technologies developed for the consortium and builds on earlier methods development performed as part of the Biomedical Informatics Research Network (BIRN (Keator et al., 2008) and the Alzheimer's Disease Neuroimaging Initiative (ADNI (The Alzheimer’s Disease Neuroimaging Initiative (ADNI): MRI Methods - Jack - 2008 - Journal of Magnetic Resonance Imaging - Wiley Online Library, n.d.)). Specifically, a standard PING scan session included: 1) a 3D T1-weighted inversion prepared RF-spoiled gradient echo scan using prospective motion correction (PROMO), for cortical and subcortical segmentation; 2) a 3D T2-weighted variable flip angle fast spin echo scan, also using PROMO, for detection and quantification of white matter lesions and segmentation of VV; 3) a high angular resolution diffusion imaging (HARDI) scan, with integrated B0 distortion correction (DISCO), for segmentation of white matter tracts and measurement of diffusion parameters; and 4) a resting state blood oxygenation leveldependent (BOLD) fMRI scan, with integrated distortion correction. Pulse sequence parameters used across (3 T) scanner manufacturers (GE, Siemens, and Phillips) and models were optimized for equivalence in contrast properties and consistency in imagederived quantitative measures (Jernigan et al., 2016).

**Pixar**

One hundred twenty-two 3.5–12-year-old children (M(s.d.) = 6.7(2.3); 64 females) participated in the study (https://openfmri.org/dataset/ds000228/). Child and adult participants were recruited from the local community. All adult participants gave written consent; parent/guardian consent and child assent was received for all child participants. Recruitment and experiment protocols were approved by the Committee on the Use of Humans as Experimental Subjects (COUHES) at the Massachusetts Institute of Technology. Whole-brain structural and functional MRI data were acquired on a 3- Tesla Siemens Tim Trio scanner located at the Athinoula A. Martinos Imaging Center at MIT. Children under age 5 years used one of two custom 32-channel phased-array head coils made for younger (n = 3, M(s.d.) = 3.91(.42) years) or older (n = 28, M(s.d.) = 4.07(.42) years) children; all other participants used the standard Siemens 32-channel head coil. T1-weighted structural images were collected in 176 interleaved sagittal slices with 1 mm isotropic voxels (GRAPPA parallel imaging, acceleration factor of 3; adult coil: FOV: 256 mm; kid coils: FOV: 192 mm). Functional data were collected with a gradient-echo EPI sequence sensitive to Blood Oxygen Level Dependent (BOLD) contrast in 32 interleaved near-axial slices aligned with the anterior/posterior commissure and covering the whole brain (EPI factor: 64; TR: 2 s, TE: 30 ms, flip angle: 90°). This data was obtained from the OpenfMRI database, accession number is ds000228. Dataset version 1.0.2 (MRI Data of 3-12 Year Old Children and Adults during Viewing of a Short Animated Film, n.d.)

**SALD**

The data was generated in the Southwest University Adult Lifespan Dataset (SALD), which comprises a large cross-sectional sample (n = 494; age range = 19-80) undergoing a multi-modal (sMRI, rs-fMRI, and behavioral). All data were collected at the Southwest University Center for Brain Imaging using a 3.0-T Siemens Trio MRI scanner (Siemens Medical, Erlangen, Ger- many). A magnetization-prepared rapid gradient echo (MPRAGE) sequence was used to acquire high-resolution T1-weighted anatomical images (repetition time=1,900ms, echo time=2.52ms, inversion time=900ms, flip angle=90 degrees, resolution matrix=256×256, slices=176, thickness =1.0mm, and voxel size=111mm3) (Wei et al., 2018).
